## Supplemental file 1 CTR20210349 for "Autologous P63+ lung progenitor cell transplantation in idiopathic pulmonary fibrosis: a phase 1 clinical trial"

| | | | |
|---|---|---|---|
| **Registration Number** | CTR20210349 | **Trial Status** | Completed |
| **Applicant** | Ting Zhang | **Initial Disclosure Date** | 2021-03-01 |
| **Sponsor Name** | Regend Therapeutics XLotus (Jiangxi) Co, Ltd. | | |

### 1. Title and Background Information

| | |
|---|---|
| **Registration Number** | CTR20210349 |
| **Related Registration Number** | NA |
| **Drug Name** | REGEND001 Autologous Therapy Product |
| **Drug Categories** | Biological products |
| **Clinical Trial Application Number** | Private |
| **Indications** | Early and mid-stage idiopathic pulmonary fibrosis (IPF) |
| **Official Title** | An Open-label Clinical Study to Explore the Safety, Tolerability and Preliminary Efficacy of REGEND001 Autologous Therapy Product for Treatment of Idiopathic Pulmonary Fibrosis (IPF). |
| **Common Title** | Exploratory Study of REGEND001 Autologous Therapy Product for the Treatment of Idiopathic Pulmonary Fibrosis (IPF) |
| **Protocol Number** | XHYX-IND-IPF-P1 |
| **Latest Version Number** | V4.0 |
| **Date of Latest Version** | 2021-05-18 |
| **Combination Therapy** | No |

### 2.Sponsor Information

| | | | |
|---|---|---|---|
| **Sponsor Name** | Regend Therapeutics XLotus (Jiangxi) Co, Ltd. | | |
| **Applicant** | Ting Zhang | **Landline Number** | 021-60646685 |
| **Mobile Number** | 18516646727 | **Email Address** | |
| **Contact Address** | Building 8, Nanchang National Pharmaceutical | **Contact Postcode** | 330096 |

| | |
|---|---|
| | International Innovation Park Joint Research Institute, 269 Aixi Lake North Road, High-tech Development Zone, Nanchang, Jiangxi Province |

### 3. Clinical Trial Information

| **(1). Trial Objective**: | |
|---|---|
| Primary Objective:<br>To evaluate the safety and tolerability of different doses of REGEND001 autologous therapy product for the treatment of idiopathic pulmonary fibrosis (IPF).<br>Secondary Objective:<br>To assess the efficacy of different doses of REGEND001 autologous therapy product for the treatment of IPF and recommend appropriate cell therapy doses for subsequent clinical studies. | |
| **(2). Trial design** | |
| **Trial Classification** | Safety and efficacy |
| **Trial Phase** | Exploratory Phase 1 clinical trial |
| **Design Type** | Single-arm trial |
| **Randomization** | Non-randomized |
| **Blinding** | Open-label |
| **Trial Scope** | Domestic trials |
| **(3). Subject Information** | |
| **Age** | 50 to 75 years old (Including ages 50 and 75) |
| **Sexes Eligible for Study** | Male and Female |
| **Accept Healthy Volunteers** | No |
| **Inclusion Criteria:** | 1). Male or female, aged between 50 to 75 (Including ages 50 and 75); |
| | 2). Subjects diagnosed with IPF according to guidelines for the diagnosis of idiopathic pulmonary fibrosis 2018 edition; |
| | 3). Subjects with 30%~79% of the predicted value in diffusing capacity for carbon monoxide (DLCO) and more than 50% of the predicted value in forced vital capacity (FVC) in pulmonary function tests 3 months before screening; |
| | 4). Subjects with typical High-resolution computed tomography (HR- |

| | |
|---|---|
| | CT) imaging findings of idiopathic pulmonary fibrosis in the past 12 months; |
| | 5). Subjects tolerant to bronchofiberscope; |
| | 6). Subjects fully informed of the purpose, method and possible discomfort of the trial, agreeing to participate in the test, and voluntarily signing the informed consent; |
| | 7). Subjects with good adherence, willingness to take medication and regular follow-up examinations as required by the protocol ； |
| | 8). Subjects able to understand and cooperate with the completion of pulmonary function tests. |
| **Exclusion Criteria:** | 1) Subjects who cannot tolerate cell therapy |
| | 2) Pregnant or lactating women; |
| | 3) Subjects with syphilis or any of human immunodeficiency virus (HIV), hepatitis B virus (HBV), hepatitis C virus (HCV) positive antibody; Of which stable HBV carriers after drug treatment (DNA titer ≤500 IU/mL or copy number <1000 copies/mL) and cured hepatitis C patients (HCV RNA is negative) can be enrolled； |
| | 4) Subjects with malignant tumors or a history of malignant tumors; |
| | 5) Subjects with taking drugs which caused lung fibroblast such as amiodarone in a long term before screening; |
| | 6) Subjects with infections in lung or other site, including bacterial and viral infections, requiring intravenous treatment before cell transplantation; |
| | 7) Subjects with a history of invasive or noninvasive mechanical ventilation within 4 weeks; |
| | 8) Subjects with any of the following lung diseases: asthma, active tuberculosis, pulmonary embolism, pneumothorax, pulmonary hypertension, pneumoconiosis, etc.; lung cancer, bronchiolitis obliterans or other active lung disease; Pneumonia currently or within the last 4 weeks; Pneumonectomy Previously; |
| | 9) Subjects needing oxygen therapy currently (oxygen therapy time> 15h/d); |
| | 10) Subjects suffering from serious other systemic diseases, such as myocardial infarction, unstable angina, liver cirrhosis, acute glomerulonephritis, connective tissue disease, etc.; |
| | 11) Subjects with following results: leukopenia (leukopenia < 4×10^9/L) or agranulocytosis (leukocyte < 1.5×10^9/L or neutrophils < 0.5×10^9/L) of any cause; Blood creatinine > 2.5 times the upper limit of normal; Alanine transaminase (ALT) and Aspartate transaminase (AST) > 2.5 times the upper limit of |

| | |
|---|---|
| | normal values in the laboratory tests. |
| | 12) Subjects with a history of mental illness or suicide risk, epilepsy or other central nervous system disorders |
| | 13) Subjects with severe arrhythmias (such as ventricular tachycardia, frequent supraventricular tachycardia, atrial fibrillation, atrial flutter, etc.) or atrioventricular block of degree II or above, shown by 12-lead Electrocardiogram (ECG); |
| | 14). Subjects with a history of abusing alcohol and illicit drug; |
| | 15). Subjects who are allergic to cattle products; |
| | 16). Subjects who participated in other clinical trials in the past 3 months; |
| | 17). Subjects with poor compliance and difficult to complete the investigation; |
| | 18). Investigators, employees of research centers or family members of them (none of whom are suitable to participate in the trial to ensure the objectivity of the research); |
| | 19). Subjects who had an acute exacerbation of IPF or hospitalized for other respiratory diseases 3 or more times in the past 1 year; |
| | 20). Subjects who take nintedanib for medication within 1 month, or plan to continue taking nintedanib for medication; |
| | 21). Subjects with other acquired or congenital immunodeficiency disorders, or with a history of organ transplantation or cell transplant therapy; |
| | 22). Subjects whose expected survival may be less than one year judged by the investigator; |
| | 23). Male participants of childbearing potential and female participants within childbearing age were reluctant to use effective contraception from the time of signing the informed consent to 6 months after cell therapy; |
| | 24). Subjects assessed as inappropriate to participate in this clinical trial by investigator. |

#### (4). Interventions

| **Investigational Drug** | **Serial Number** | **Name** | **Administration** |
|---|---|---|---|
| | 1). | English Generic Name：REGEND001 Autologous Therapy Product | Dosage Form：Intratracheal administration preparation<br>Specifications：14mL/bag<br>Administration：Intratracheal administration through fiberoptic bronchoscopy |

| | | | |
|---|---|---|---|
| | | | Administration Frequency：Single dose |
| **Control Drug** | **Serial Number** | **Name** | **Administration** |
| | | NA | NA |

**(5). Outcome Measures**

| **Primary Outcome and Assessment Time** | **Serial Number** | **Indicators** | **Assessment Time** | **Type of indicators** |
|---|---|---|---|---|
| | 1) | Incidence and severity of the cell therapy-related adverse events (AEs) | Within 24 weeks after treatment | Safety Indicator |

| **Secondary Outcomes and Assessment Time** | **Serial Number** | **Indicators** | **Assessment Time** | **Type of indicators** |
|---|---|---|---|---|
| | 1) | Incidence of complication related to bronchoscopy； | Within 24 weeks after treatment | Safety Indicator |
| | 2) | Change of tumor markers from baseline | 12 and 24 weeks after treatment | Safety Indicator |
| | 3) | Change of routine safety assessments (Blood routine, Urine routine, Blood biochemistry, 12-lead Electrocardiogram (ECG)) | Within 24 weeks after treatment | Safety Indicator |
| | 4) | Change of the percentage of predicted value for single-breath diffusing capacity for carbon monoxide (DLCO-sb) from baseline | 4, 12 and 24 weeks after treatment | Efficacy Indicator |
| | 5) | Change of forced vital capacity (FVC) from baseline | 4, 12 and 24 weeks after treatment | Efficacy Indicator |
| | 6) | Change of the ratio of diffusing capacity for carbon monoxide/ the alveolar volume | 4, 12 and 24 weeks after treatment | Efficacy Indicator |

| | | | | |
|---|---|---|---|---|
| | | (DLCO/VA) from baseline | | |
| | 7) | Change of 6-minute-walk test (6MWT) from baseline | 4, 12 and 24 weeks after treatment | Efficacy Indicator |
| | 8) | Change of St. George's respiratory questionnaire (SGRQ) scale from baseline | 4, 12 and 24 weeks after treatment | Efficacy Indicator |
| | 9) | Change of imaging of lung by high resolution computed tomography (HR-CT) | 24 weeks after treatment | Efficacy Indicator |
| | 10) | Idiopathic pulmonary fibrosis (IPF) exacerbation events (Frequency and severity) | Within 24 weeks after treatment | Efficacy Indicator |
| **(6). Has Data Monitoring Committee（DMC）** | No | | | |
| **(7). Trial injury insurance for subjects** | Yes | | | |

#### 4. Investigator Information

| **(1). Principal Investigator Information** | | | |
|---|---|---|---|
| **Name** | Zuojun Xu | **Degree** | Medical Doctor (MD) |
| **Professional Title** | Senior | **Phone Number** | 010-69156114 |
| **Email** | | **Postal Address** | No. 1, Shuaifu Garden, Dongcheng District, Beijing, Beijing Municipality |
| **Postal Code** | 100730 | **Institution Name** | Peking Union Medical College Hospital |

| **(2). Institution Information** | | | | |
|---|---|---|---|---|
| **Serial Number** | **Institution Name** | **Principal Investigator** | **Country or Region** | **Province (State) - City** |

| | | | | |
|---|---|---|---|---|
| 1) | Peking Union Medical College Hospital | Zuojun Xu | China | Beijing Municipality - Beijing City |
| 2) | Ruijin Hospital, Shanghai Jiao Tong University School of Medicine l | Jieming Qu | China | Shanghai Municipality - Shanghai City |
| 3) | The First Affiliated Hospital of Guangzhou Medical University | Qun Luo | China | Guangdong Province - Guangzhou City |

#### 5. Ethics Committee Information

| **Serial Number** | **Name** | **Review Conclusion** | **Approval Date/Review Date** |
|---|---|---|---|
| 1) | Ethics Committee for Clinical Trials of Drugs at Peking Union Medical College Hospital Chinese Academy of Medical Sciences | Agreed after modification | 2021-01-28 |
| 2) | Rapid review approval from the Ethics Committee for Clinical Trials of Drugs at Peking Union Medical College Hospital Chinese Academy of Medical Sciences | Agree | 2021-06-07 |

#### 6. Trial Status

| **(1). Trial Status** | |
|---|---|
| Completed | |
| **(2). Participant Enrollment** | |
| **Target Enrollment** | Domestic: 24 participants； |
| **Enrolled Participants** | Domestic: 12 participants； |
| **Actual Total Enrollment** | Domestic: 12 participants； |
| **(3). Participant Enrollment and Study Completion Date** | |
| **Date of First Subject Signing Informed Consent** | Domestic：2021-05-14； |
| **Date of First Subject Enrollment** | Domestic：2021-07-19； |
| **Study Completion Date** | Domestic：2023-06-09； |
