## Supplemental file 2 Ethics Committee Approval (English Version) for "Autologous P63+ lung progenitor cell transplantation in idiopathic pulmonary fibrosis: a phase 1 clinical trial"

### Ethics Committee Approval for Expedited Review from the Drug Clinical Trial Ethics Committee of Peking Union Medical College Hospital, Chinese Academy of Medical Sciences

Project Number: 002401

| | | | |
|---|---|---|---|
| Drug/Medical Device Name | REGEND001 Autologous Therapy Product | Registration Category | Biological Products Class 1 |
| Applicant | Regend Therapeutics XLotus (Jiangxi) Co, Ltd. | Task Source | NMPA |
| Professional Group | Department of Respiratory Medicine | Application Matter | Domestic Drug Registration |
| Approval Number | CXSL1900019 | Principal Investigator | Zuojun Xu |
| Chairperson | Liying Cui | Vice Chairpersons | Xiaomei Zhai, Hua Bai |
| Protocol Title | Protocol for An Open-Label Clinical Study to Explore the Safety, Tolerability, and Preliminary Efficacy of REGEND001 Autologous Therapy Product for Treatment of Idiopathic Pulmonary Fibrosis (IPF) | | |
| Protocol Number | XHYX-IND-IPF-P1 | | |

This research project was approved by the Drug Clinical Trial Ethics Committee of Peking Union Medical College Hospital, Chinese Academy of Medical Sciences on January 28, 2021. Approval number: KS2021039. Approval Comment: Approved.

Review Comment:

☑ Approved ☐ Requires Committee Review ☐ Disapproved

Frequency of Follow-Up Review:

☑ 3 months ☐ 6 months ☐ 1 year ☐ None ☐ Other

Signature of the Chairperson/Vice Chairperson of the Ethics Committee: Liying Cui

Date: 2021-06-07

*The materials for this approval are attached.

Attachment:

1. Protocol (Version No.: 4.0; Version Date: May 18, 2021)
2. Informed Consent Form (Version No.: 2.0; Version Date: May 19, 2021)
3. Recruitment Advertisement (Version No.: 2.0; Version Date: May 19, 2021)
4. eCRF (Version No.: 1.0; Version Date: April 30, 2021)
5. eCRF (Version No.: 1.1; Version Date: May 27, 2021)
6. Document Amendment Instructions (Protocol, Informed Consent, Recruitment Advertisement, eCRF)
7. Interim Diagnosis Form (Version No.: A/0)
8. Subject Inclusion/Exclusion Criteria Determination Form (Version No.: A/0)
9. 6-Minute Walking Distance Test Record Form (Version No.: A/0)
10. Medical Research Council Dyspnea Scale (Version No.: A/0)
11. Saint George's Respiratory Questionnaire (Version No.: A/0)
12. Bronchial Basal Cell Collection Record Form (Version No.: A/0)
13. Bronchial Basal Cell Transportation Receipt Form (Version No.: A/0)
14. Cell Preparation Distribution Application Form (Version No.: A/0)
15. Cell Preparation Transportation Receipt Form (Version No.: A/0)
16. Cell Preparation Infusion Record Form (Version No.: A/0)
17. Cell Preparation Return Registration Form (Version No.: A/0)
18. Document Amendment Instructions (Procedural Forms)

1. This Ethics Committee is independent and complies with ICH GCP, China GCP, and relevant local regulations. All attending members are within their valid term of office.
2. This Ethics Committee will keep the clinical research materials and related content confidential. Additionally, there is no conflict of interest with this research project.
3. The approval is valid for one year. Please report the clinical trial status in a timely manner according to the corresponding follow-up review frequency.
4. Address of the Ethics Committee: 41 Damucang Hutong, Xicheng District, Beijing. Contact person: Jiali Tian, Phone number: 010-6915839

Drug Clinical Trial Ethics Committee of Peking Union Medical College Hospital, Chinese Academy of Medical Sciences
