## Supplemental file 3 Informed consent forms (English Version) for "Autologous P63+ lung progenitor cell transplantation in idiopathic pulmonary fibrosis: a phase 1 clinical trial"

### Informed Consent Form · Information Page

Participant Information

Dear Sir/Madam:

We invite you to participate in an open-label clinical study titled " Explore the Safety, Tolerability, and Preliminary Efficacy of REGEND001 Autologous Therapy Product for Treatment of Idiopathic Pulmonary Fibrosis (IPF)”. This study has been approved by the National Medical Products Administration and the ethics committee of the hospital. Before deciding whether to participate, please read the following information carefully, as it will help you understand the purpose, procedures, duration of the study, and potential benefits, risks, and discomforts of participation. You are encouraged to discuss it with your family or friends to help make your decision. If you have any questions, please consult your responsible doctor. Once all your questions are answered, you are satisfied with the explanations about this study, and you decide to participate, you will be asked to sign this informed consent form. Participation in the study is voluntary, you can agree to participate or not.

The following is an introduction to this study:

#### 1. Research Drug Information

REGEND001 autologous therapy product (referred to as REGEND001 cells) is developed by Regend Therapeutics XLotus (Jiangxi) Co, Ltd. for the treatment of idiopathic pulmonary fibrosis. Idiopathic pulmonary fibrosis is a chronic progressive fibrotic interstitial pneumonia of unknown etiology. Its main manifestations include progressively worsening dyspnea, restrictive ventilatory impairment, and gas exchange disorders, ultimately leading to hypoxemia and even respiratory failure. Traditional treatments for idiopathic pulmonary fibrosis mainly involve the use of anti-inflammatory and anti-fibrotic medications. Currently approved treatments in China include pirfenidone and nintedanib, which can slow down the decline in lung function to some extent but are difficult to improve lung function or reverse the disease course, and cannot effectively prevent fibrosis. Currently, apart from performing whole lung transplantation surgery for idiopathic pulmonary fibrosis, there is a lack of effective conventional treatment worldwide, primarily due to the lack of effective methods for regenerating and repairing damaged alveolar structures in the lungs.

The preparation of REGEND001 cells requires the research physician to obtain a small amount of bronchial brush tissue via bronchoscopic brushing when your condition is stable. Subsequently, using the unique technology of Regend Therapeutics XLotus (Jiangxi) Co, Ltd. (the sponsor), bronchial basal cells are isolated from the brush tissue and expanded in vitro to prepare the REGEND001 autologous therapy product. After injection into the bronchial airway, REGEND001 cells migrate from the injection site to the damaged area of the distal airway in a radiating distribution pattern, specifically settling in damaged lung tissue. Once settled, REGEND001 cells undergo normal metabolism similar to native lung stem cells, primarily participating in the repair of lung alveoli. REGEND001 cells do not migrate to other organs outside the lungs (such as liver, kidney, spleen, etc.). In undamaged lung tissue, transplanted cells cannot persist for a long term. Currently, the production process of Regend Therapeutics XLotus (Jiangxi) Co, Ltd. has been standardized and matured, and the preparation of this drug can be verified. With the support and monitoring of established strict quality assurance and quality control systems, the isolation, expansion, and preparation of bronchial basal cells can be efficiently carried out to produce REGEND001 autologous therapy product that meet clinical quality standards. This drug has not yet been approved for formal market release by regulatory authorities.

#### 2. Research Information

##### 2.1 Study Objectives

(1) Primary Objective: To evaluate the safety and tolerability of different doses of REGEND001 autologous therapy product for the treatment of idiopathic pulmonary fibrosis (IPF).

(2) Secondary Objective: To assess the efficacy of different doses of REGEND001 autologous therapy product for IPF and recommend appropriate cell therapy doses for subsequent clinical studies.

##### 2.2 Treatment Groups, Number of Participants, and Study Duration

This is a multi-center, open-label, dose-escalation, exploratory clinical study. It plans to include 15-24 participants with idiopathic pulmonary fibrosis and divide them into single-dose groups (Group 1: 3-6 participants, 0.6 times the standard dose; Group 2: 3-6 participants, 1 time the standard dose; Group 3: 3-6 participants, 2 times the standard dose; Group 4: 6 participants, 3.3 times the standard dose). The standard dose is $1x10^6$ cells/kg/person. The study includes a screening period (2 weeks), cell preparation period (4-8 weeks), reinfusion therapy, and post-treatment follow-up (24 weeks). Long-term follow-up will occur at 1, 2, 3, 5, 8, and 10 years post treatment, and then every 5 years until your death or loss to follow-up. Follow-up indicators include lung function, high-resolution chest CT and survival status.

**2.3 Who is Eligible to Participate?**

You must meet all of the following criteria to enter the study:

1. Male or female, aged between 50 to 75 (Including ages 50 and 75);
2. Subjects diagnosed with IPF according to guidelines for the diagnosis of idiopathic pulmonary fibrosis 2018 edition;
3. Subjects with 30%~79% of the predicted value in diffusing capacity for carbon monoxide (DLCO) and more than 50% of the predicted value in forced vital capacity (FVC) in pulmonary function tests 3 months before screening;
4. Subjects with typical High-resolution computed tomography (HR-CT) imaging findings of idiopathic pulmonary fibrosis in the past 12 months;
5. Subjects tolerant to bronchofiberscope;
6. Subjects fully informed of the purpose, method and possible discomfort of the trial, agreeing to participate in the test, and voluntarily signing the informed consent;
7. Subjects with good adherence, willingness to take medication and regular follow-up examinations as required by the protocol；
8. Subjects able to understand and cooperate with the completion of pulmonary function tests.

**2.4 Who Is Not Suitable to Participate in This Study**

If you have any of the following characteristics, you will not be suitable to participate in this study:

1. Subjects who cannot tolerate cell therapy

2. Pregnant or lactating women;
3. Subjects with syphilis or any of human immunodeficiency virus (HIV), hepatitis B virus (HBV), hepatitis C virus (HCV) positive antibody; Of which stable HBV carriers after drug treatment and cured hepatitis C patients can be enrolled;
4. Subjects with malignant tumors or a history of malignant tumors;
5. Subjects with taking drugs which caused lung fibroblast such as amiodarone in a long term before screening;
6. Subjects with infections in lung or other site, including bacterial and viral infections, requiring intravenous treatment before cell transplantation;
7. Subjects with a history of mechanical ventilation within 4 weeks;
8. Subjects with any of the following lung diseases: asthma, active tuberculosis, pulmonary embolism, pneumothorax, pulmonary hypertension, pneumoconiosis, etc.; lung cancer, bronchiolitis obliterans or other active lung disease; Pneumonia currently or within the last 4 weeks; Pneumonectomy Previously;
9. Subjects needing oxygen therapy currently (oxygen therapy time> 15h/d);
10. Subjects suffering from serious other systemic diseases, such as myocardial infarction, unstable angina, liver cirrhosis, acute glomerulonephritis, connective tissue disease, etc.;
11. Subjects with following results : leukopenia or agranulocytosis of any cause; Blood creatinine > 2.5 times the upper limit of normal; Alanine transaminase (ALT) and Aspartate transaminase (AST) > 2.5 times the upper limit of normal values in the laboratory tests.
12. Subjects with a history of mental illness or suicide risk, epilepsy or other central nervous system disorders
13. Subjects with severe arrhythmias (such as ventricular tachycardia, frequent supraventricular tachycardia, atrial fibrillation, atrial flutter, etc.) or atrioventricular block of degree II or above, shown by 12-lead Electrocardiogram (ECG);
14. Subjects with a history of abusing alcohol and illicit drug;
15. Subjects who are allergic to cattle products;
16. Subjects who participated in other clinical trials in the past 3 months;

17. Subjects with poor compliance and difficult to complete the investigation;
18. Investigators, employees of research centers or family members of them (none of whom are suitable to participate in the trial to ensure the objectivity of the research);
19. Subjects who had an acute exacerbation of IPF or hospitalized for other respiratory diseases 3 or more times in the past 1 year;
20. Subjects who take nintedanib for medication within 1 month, or plan to continue taking nintedanib for medication;
21. Subjects with other acquired or congenital immunodeficiency disorders, or with a history of organ transplantation or cell transplant therapy;
22. Subjects whose expected survival may be less than one year judged by the investigator;
23. Male participants of childbearing potential and female participants within childbearing age were reluctant to use effective contraception from the time of signing the informed consent to 6 months after cell therapy;
24. Subjects assessed as inappropriate to participate in this clinical trial by investigator.

**2.5 Related Examinations and Possible Costs**

During your participation in this study, you will be required to undergo relevant safety and efficacy examinations as per the protocol (including laboratory tests, imaging examinations, etc., details can be found in the study protocol below), to enable the research doctors to better monitor your condition. The investigational drug in this study is provided at no cost to you, and all study visits, laboratory tests, and procedures associated with the study are free of charge. However, regular medical examinations required for your routine healthcare needs (such as treatment and examinations for concurrent illnesses) are not covered under this study. Regend Therapeutics XLotus (Jiangxi) Co, Ltd. will not be responsible for paying for unrelated treatments or examinations.

**2.6 Study Procedure**

This study is divided into several phases: screening period, cell collection period, baseline period, transplantation treatment period, and follow-up observation period. Each phase will be explained in detail below. Throughout the clinical trial, researchers will assess your condition, evaluate safety events and inquire about other medication usage.

Upon signing the informed consent form, you will enter the screening period (14 days). To determine your eligibility for the study, the research doctor will collect demographic information (such as age, gender, ethnicity), medical history, and conduct a series of examinations. These include physical examination, vital signs, blood routine, urine routine, pregnancy test for women, blood biochemistry, coagulation profile, cardiac enzyme spectrum, autoimmune disease antibody testing, tumor markers, syphilis antibody (TP-Ab), human immunodeficiency virus (HIV) antibody, hepatitis B surface antigen (HBsAg), hepatitis C virus (HCV) antibody, 12-lead electrocardiogram, high-resolution chest CT, lung function tests (ventilation + diffusion) and dyspnea scoring.

If the research doctor determines you are suitable for the study, you will proceed to the collection period (1 day) for cell collection. Before cell collection, you will undergo a physical examination, vital signs check and dyspnea scoring, to assess your disease status. Cells will be collected via bronchoscopy for preparation of the investigational drug, which may take approximately 4-8 weeks. If during this period you develop conditions that make you unsuitable for inclusion, you will not be able to continue in the study. After bronchoscopic cell sampling, you will be observed for about 2 hours. The duration of safety observation may be extended based on your safety condition as assessed by the research doctor.

Once the investigational drug preparation is completed, you will be notified to undergo baseline assessments at the hospital (0-5 days). This includes vital signs, physical examination, blood routine, urine routine, pregnancy test for women, blood biochemistry, blood gas analysis, coagulation profile, 12-lead electrocardiogram, lung function tests (ventilation + diffusion), assessment of 6-minute walkinging distance, dyspnea scoring and completion of the St. George's Respiratory Questionnaire.

After the research doctor confirms you are eligible for the investigational drug infusion, you will proceed to the administration period (1 day). Following vital signs and physical examination, you will receive the investigational drug infusion. After cell transplantation, you will be observed at the outpatient clinic or hospital for 2-4 hours. The researcher will decide your discharge time based on your safety and tolerance during observation.

After completion of the drug infusion, you will enter the follow-up phase:

1. 24 hours post-treatment: Vital signs and physical examination.
2. 1-week post-treatment: Vital signs, physical examination, blood routine, urine routine, blood biochemistry, 12-lead electrocardiogram.
3. 4-weeks post-treatment: Vital signs, physical examination, blood routine, urine routine, blood biochemistry, 12-lead electrocardiogram, lung function tests (ventilation + diffusion), assessment of 6-minute walking distance, completion of the St. George's Respiratory Questionnaire and dyspnea scoring.
4. 12-weeks post-treatment: Vital signs, physical examination, blood routine, urine routine, blood biochemistry, blood gas analysis, coagulation profile, lung tumor markers, high-resolution chest CT, lung function tests (ventilation + diffusion), assessment of 6-minute walking distance, completion of the St. George's Respiratory Questionnaire and dyspnea scoring.
5. 24-weeks post-treatment: Vital signs, physical examination, blood routine, urine routine, pregnancy test for women, blood biochemistry, blood gas analysis, coagulation profile, cardiac enzyme spectrum, autoimmune disease antibody testing, syphilis antibody (TP-Ab), human immunodeficiency virus (HIV) antibody, hepatitis B surface antigen (HBsAg), hepatitis C virus (HCV) antibody, lung tumor markers, high-resolution chest CT, 12-lead electrocardiogram, lung function tests (ventilation + diffusion), assessment of 6-minute walking distance, completion of the St. George's Respiratory Questionnaire and dyspnea scoring.

**2.7 Specimen Collection in the Study**

During your participation in this study, three types of specimens will be collected: blood, urine, and lung basal cells. All specimen collections will take place at the hospital where you are participating in the study. (1) Blood and Urine: These samples will be used for blood routine, blood biochemistry, coagulation function, urine routine, and other required tests in the study protocol. Throughout your participation in the trial, approximately 124.5 milliliters of blood and 60 milliliters of urine will be collected. After each collection, the blood and urine samples will be destroyed after completion of testing at the hospital. (2) Lung Basal Cells: Approximately 1 milliliter of your lung basal cells will be collected and transported to the preparation center of Regend Therapeutics XLotus (Jiangxi) Co, Ltd. for the preparation of REGEND001 autologous therapy product. If the preparation is successful, the REGEND001 autologous therapy product will be infused back into your lungs, and any remaining portion required by regulations will be stored for retesting at Regend Therapeutics XLotus (Jiangxi) Co, Ltd. for up to 30 years before being uniformly destroyed. In case of unsuccessful preparation, the specimen will be immediately destroyed.

#### 3. What do you need to do in this study?

Truthfully inform the research doctor about your health information, current treatments, and any other therapies you've started during the study period. If you withhold this information, your health may be harmed.

Strictly follow medical advice during the study period. Nintedanib must not be used during the study.

Adhere to the schedule of doctor for tests, treatments, and follow-ups. If you cannot attend scheduled appointments due to timing, financial, or other special issues, promptly contact the doctor to coordinate.

During the study period, you and your spouse should avoid pregnancy.

#### 4. Pregnancy and Breastfeeding

Women who are pregnant or breastfeeding cannot participate in this study. From the time of signing the informed consent to 6 months after receiving cell therapy, effective contraception measures must be taken. If you or your partner become pregnant during the trial period, inform the research doctor immediately. Any female participant who becomes pregnant during the study must withdraw from the trial and immediately stop using the study drug.

#### 5. Potential Benefits of Participating in this Study

Participating in this study may benefit you by potentially improving your condition. It may also provide benefits to other patients suffering from similar conditions in the future.

Although there is evidence suggesting varying degrees of improvement in participants' conditions, this does not guarantee definite effectiveness for you. If you believe this method is ineffective for your condition, consult with your doctor about other possible treatment options.

#### 6. Possible risks

**Regarding Bronchoscopy:**

Bronchoscopy is an invasive examination procedure. Patients may experience anxiety during the procedure, and they may encounter various adverse reactions such as nausea, breathlessness and coughing. Additionally, it can potentially cause damage to the respiratory mucosa.

Common complications of bronchoscopy include anesthesia accidents, bleeding, pneumothorax, mediastinal emphysema, laryngospasm or laryngeal edema, arrhythmia, cardiac arrest, severe bronchospasm, hypoxemia, infection, postoperative fever and other unforeseen events. Although bronchoscopy techniques are mature and relatively safe in clinical practice, these risks are still possible. This study involves only local segmental lavage and injection of the lung.

**Regarding Anesthesia:**

(1) Allergic reactions to anesthesia drugs: Prior to anesthesia, a small amount of medication sprayed into the throat may cause allergic reactions.

(2) Insufficient or excessive anesthesia: Inadequate local anesthesia can lead to coughing, vomiting, discomfort for the subject and increase the risk of mucosal damage during bronchoscopy. Excessive local anesthesia may cause cardiac toxicity, local tissue reactions, neurological reactions, cytotoxic reactions and systemic adverse reactions such as allergic reactions, neurotoxic reactions (numbness of lips and tongue, headache, dizziness, blurred vision, difficulty focusing or eye tremors, unclear speech, muscle twitching, incoherent speech, altered consciousness, seizures, etc.).

(3) Anesthesia accidents: After anesthesia, there may be fainting, toxicity, hematoma, nerve damage, infection at the injection site and temporary blurred vision or temporary facial paralysis.

**Regarding REGEND001 Cell Therapy:**

(1) Multi-organ dysfunction syndrome (MODS): MODS is a clinical syndrome characterized by simultaneous or sequential dysfunction or failure of two or more organs due to severe pathologic damage such as severe infection, trauma, shock and severe pancreatitis. The damaged organs include lungs, kidneys, liver, gastrointestinal tract, heart,

brain, etc., and may involve coagulation and metabolic dysfunction. Because the lung capillary network is rich, REGEND001 cells may enter the bloodstream through the lung capillary network and reach multiple organs in the body. If a large number of cells enter the bloodstream, it may further lead to organ dysfunction.

(2) Local tumor formation: Refers mainly to tumors appearing in the bronchial airway, most likely due to cell accumulation, primarily from natural metabolic apoptosis. No tumorigenicity of REGEND001 cells has been observed in animal experiments or in previous clinical studies of REGEND001 cells.

(3) Other possible complications: No clinically relevant complications were observed in previous clinical studies of REGEND001 cells. However, during this study, your condition will be closely monitored by the study physician to detect and treat any potential complications.

(4) Failure to receive treatment due to drug preparation failure: If during the preparation process it is found that cells cannot be cultured or there are abnormalities affecting cell quality, arrangements may be made to collect your lung basal cells again for re-preparation. If re-collection fails, you will no longer be able to continue participating in this study.

**Regarding Planned Examinations in the Protocol:**

(1) CT scan: Carries a certain amount of radiation with long-term carcinogenic risks.

(2) Pulmonary function test: Used to assess respiratory function, may cause coughing, chest tightness, rapid breathing, wheezing, hoarseness, throat discomfort, dizziness, and headache. These symptoms generally resolve after rest. Excessive exertion may lead to increased blood pressure, cardiovascular accidents: hyperventilation or breath-holding causing fainting; arrhythmias, myocardial infarction, hypotension, and even shock.

(3) 6-minute walking test: A functional exercise test for patients with moderate to severe cardiopulmonary diseases, may be terminated if the patient experiences chest pain, intolerable dyspnea, lower limb spasms, unsteady gait, sweating, pallor, etc.

(4) Blood draw: Some patients may experience fainting or needle phobia. After blood draw, there may be local hematoma or subcutaneous ecchymosis.

During this period, you may experience acute exacerbation of illness. Acute exacerbation of idiopathic pulmonary fibrosis refers to a sudden worsening of symptoms, increased dyspnea and decreased pulmonary function without clear triggers, leading to respiratory failure or even death.

If you experience any discomfort, new changes in your condition, or any unexpected events during the study, regardless of whether they are related to the study, please inform your physician promptly. Your physician will assess the situation and provide appropriate medical treatment.

#### 7. Regarding Fees and Compensation

After signing the informed consent form, the drug treatment, examination fees related to this trial, and treatment costs for adverse events related to this trial in your diagnostic and treatment process are all free of charge. Costs unrelated to the trial (such as other underlying diseases you may have) are your responsibility. If you participate in this study, the sponsor will provide a transportation subsidy of 200 RMB per visit. Upon completion of the study, you will receive a compensation of 1800 RMB for transportation expenses, which will be paid in cash or transferred to you in 9 installments based on the number of scheduled visits during the treatment period. If you do not complete the study as required by the protocol, you will receive compensation proportionate to the completed trial activities. The final settlement will be based on the actual number of visits.

#### 8. Participant Insurance

If you suffer any injury or damage related to the study during the treatment period, compensation will be made in accordance with relevant national laws and regulations. The sponsor will purchase insurance for this trial. If any injury related to the trial occurs, the sponsor will cover treatment costs and corresponding compensation (excluding medical accidents).

#### 9. Confidentiality of Information

Your medical records will be kept confidential within the limits permitted by law. Your medical records (including original medical records/case report forms, physical examination reports, etc.) will be kept complete and stored as required by the hospital. Researchers, sponsor representatives, ethics committees and drug regulatory authorities will be allowed to access your medical records. Any public report on the results of this study will not disclose your identity. We will make every effort to protect the privacy of your personal medical information within the limits allowed. If the research results are publicly published, your identity will not be disclosed. By signing this document, you authorize the use of your medical records as described above.

#### 10. Voluntary Participation and Withdrawal

Participation in the study is entirely voluntary based on your willingness. You may refuse to participate in this study or withdraw from the study at any time without affecting your relationship with the doctor or any loss to your medical care or other benefits.

For your best interest, the doctor or researcher may suspend your participation in the study at any time.

We will request your withdrawal from the study under the following circumstances:

(1) Certain test results indicate that you are unsuitable for participating in this study;

(2) You cannot cooperate with treatment or attend follow-up visits promptly;

(3) New health issues arise during the study period;

(4) Pregnancy or deciding to become pregnant;

(5) For your benefit, the study doctor believes that the study should be stopped.

If you withdraw from the study midway, the data collected up to that point will still be used for the study's result analysis. Additionally, for your safety, tests will be conducted on the day of withdrawal, and the tests will be the same as the 24-week post-treatment examinations.

#### 11. Information, Consultation, and Contact Information

You may ask any questions about this study at any time and receive corresponding answers.

If there is any important new information during the study that may affect your willingness to continue participating, your doctor will inform you promptly, and if necessary, obtain your informed consent again.

If you have any questions about this study, you can consult your study doctor. If you have any concerns about your rights in participating in this study, you may consult the Ethics Committee of our institution at the following contact number: 010-69158355.

### Informed Consent Form · Signature Page

Study Title: An Open-Label Clinical Study to Explore the Safety, Tolerance, and Preliminary Efficacy of REGEND001 Autologous Therapy Product For the Treatment of Idiopathic Pulmonary Fibrosis (IPF)

Sponsor: Regend Therapeutics XLotus (Jiangxi) Co, Ltd.

Clinical Trial Approval Number from National Medical Products Administration: 2020LP00065

**Subject's Declaration of Consent**

I (the subject) have read the information regarding the study mentioned above, and have had the opportunity to discuss and ask questions about this study with the doctor. All my questions have been satisfactorily answered.

I understand the risks and benefits associated with participating in this study. I acknowledge that participation is voluntary, and I confirm that I have had sufficient time to consider this decision. I understand:

I can consult the doctor for more information at any time.

I can withdraw from this study at any time without discrimination or retaliation, and my medical treatment and rights will not be affected.

If I do not follow the study plan or if there are reasons related to injury or other circumstances that make continued participation inappropriate, the study doctor may terminate my participation.

If I withdraw from the study, I will inform the doctor of changes in my condition and complete the necessary examinations.

If I need any other treatment due to changes in my condition, I will seek the doctor's advice beforehand or inform the doctor truthfully afterwards.

I agree that the hospital's supervisory department, ethics committee, or researchers may access my research data and use it for academic exchanges and other purposes.

I will receive a copy of the signed and dated Informed Consent Form (including the Informed Consent Information Page and Signature Page). I decide to consent to participate in this study and will follow medical advice. I also agree to undergo safety follow-ups after participating in the study.

Subject's Name (in block letters):

Subject's Signature:

Contact Phone:

Date: Year Month Day Hour Minute

Legal Guardian's Name (in block letters):

Legal Guardian's Signature:

Contact Phone:

Date: Year Month Day Hour Minute

**Doctor's Declaration**

I confirm that I have explained the details of this study to the subject, including their rights and the potential benefits and risks, and have provided them with a signed copy of the Informed Consent Form.

Researcher's Name (in block letters):

Researcher's Signature:

Contact Phone:

Date: Year Month Day Hour Minute
