## Supplemental file4 Protocol for IPF Clinical Trial for "Autologous P63+ lung progenitor cell transplantation in idiopathic pulmonary fibrosis: a phase 1 clinical trial"

### Protocol for An Open-label Clinical Study to Explore the Safety, Tolerability and Preliminary Efficacy of REGEND001 Autologous Therapy Product for Treatment of Idiopathic Pulmonary Fibrosis (IPF)

Protocol Version Number: V4.0

Version Date: May 18th, 2021

Research Center: Peking Union Medical College Hospital; The First Affiliated Hospital of Guangzhou Medical University; Ruijin Hospital, Shanghai Jiaotong University School of Medicine

Principal Investigator: Zuojun Xu

Sponsor: Regend Therapeutics XLotus (Jiangxi) Co, Ltd.

**Confidentiality Statement**

The information contained in this research protocol is provided only to the researcher, ethics committees, and relevant institutions of this project for reviewing. The relevant information contained in this research protocol involves commercial confidentiality and commercial information, this business information is confidential or proprietary; unless required by relevant laws and regulations, no one shall disclose these confidential contents without authorization. Any individual or enterprise who has access to the file should understand the propriety or confidentiality of the file, and shall not disclose the file without authorization. The above restrictions on the confidentiality of documents will apply to any materials that are marked as proprietary or confidential in the future.

### 1 RESEARCH PURPOSE

Primary purpose: To evaluate the safety and tolerability of different doses of REGEND001 autologous therapy product for the treatment of idiopathic pulmonary fibrosis (IPF).

Secondary purpose: To assess the efficacy of different doses of REGEND001 autologous therapy product for the treatment of IPF and recommend appropriate cell therapy doses for subsequent clinical studies.

#### 1.1 STUDY DESIGN

Multicenter, open-label, dose-escalation, exploratory clinical research.

#### 1.2 OVERALL DESIGN

This study is an open-label clinical research on the safety, tolerance, and primary efficacy of REGEND001 autologous therapy product treating idiopathic pulmonary fibrosis, with a follow-up period of 24 weeks.

#### 1.3 DOSE-ESCALATION GROUPS

The standard dose of REGEND001 autologous therapy product is $1\times10^6$ cells /kg bodyweight /person.

Each dose group will enroll around 3-6 patients. Before starting the next dose group, safety evaluation must be done for 3 patients of the current dose group at 4 weeks post-transplantation. If 1 patient from a certain dose group undergo adverse events that would terminate the study within the evaluation window (28 days after receiving REGEND001 autologous therapy product), 3 more patients should be enrolled for this dose group; the next dose group cannot start unless none of the new enrolled 3 patients undergo adverse events that would terminate the study.

The first group with 3-6 patients, receiving a single administration at 0.6 times standard dose;

The second group with 3-6 patients, receiving a single administration at 1 times standard dose (the median safe dose in the previous clinical trial);

The third group with 3-6 patients, receiving a single administration at 2 times standard dose;

The fourth group with 6 patients, receiving a single administration at 3.3 times standard dose;

The following doses will be arranged according to the safety measures of these 4 dose groups.

#### 1.4 STUDY PROCEDURE

This study includes screening stage V1, cell collection stage V2, baseline stage V3 (0-5 days before cell transplantation), REGEND001 autologous therapy product transplantation stage V4, follow-up stage V5-V9, safety follow-up stage after research. Follow-up stage includes: V5 (24 hours ± 4 hours after transplantation), V6 (1 week ± 2 days after transplantation), V7 (4 weeks ± 3 days after transplantation), V8 (12 weeks ± 5 days after transplantation), V9 (24 weeks ± 7 days after transplantation). During the study, the vital signs, laboratory examinations, 12-lead electrocardiogram, chest HRCT, pulmonary functions, 6-minute walking distance, questionnaires, concomitant medications, adverse events, etc. will be checked and recorded.

Screening stage V1: After the patient signing the informed consent, relevant physical examination and check of previous physical examination will be done. The investigator will check inclusion and exclusion criteria, making judgement on whether the patient is eligible.

Cell collection stage V2: Tissue collection through bronchoscopy, cell isolation and expansion of around 4-8 weeks before autologous transplantation of REGEND001 autologous therapy product.

Baseline stage V3 (0-5 days before cell transplantation): 4-8 weeks later, conduct relevant examinations again and check inclusion and exclusion criteria. For idiopathic pulmonary fibrosis patients who are still eligible, they can formally join in this clinical trial and will receive autologous transplantation of REGEND001 cell products.

Cell transplantation stage V4 (the day of cell transplantation): According to the dose group each patient was assigned, a single administration of REGEND001 cell products at the corresponding dose were autologously transplanted to different lung segments through bronchoscopy. Investigator should be on site during the transplantation, confirming the dose, starting and stopping time of the transplantation, and recording correctly. Close observation of the patients in the outpatient or hospital for 2-4 hours after administration, and the investigator will determine when the patients can leave based on the tolerance of the patients. After leaving the hospital, the patients should return back to the research center as required for follow-up.

Follow-up stage: The patients will stop receiving REGEND001 during the follow-up stage, and continue with usual treatment which can be adjusted according to the doctor’s orders.

Each patient will be followed up for 24 weeks after the administration, with a total of 5 visits. The follow-up time points including V5 (24 hours ± 4 hours after transplantation), V6 (1 week ± 2 days after transplantation), V7 (4 weeks ± 3 days after transplantation), V8 (12 weeks ± 5 days after transplantation), and V9 (24 weeks ± 7 days after transplantation). During the study, check and record the vital signs, laboratory examinations, 12-lead electrocardiogram, chest HRCT, pulmonary functions, 6-minute walking distance, questionnaires, concomitant medications, adverse events, etc.

If the patients undergo adverse events/severe adverse events, the situation should be documented in detail and followed up until the outcome is clear.

Safety follow-up stage after research: After the end of the follow-up stage in this study, the patient will enter the post-study safety follow-up until the patient's death or lost. The patient will be followed up once each at 1, 2, 3, 5, 8, and 10 years after treatment, and then once every five years.

### Study Flowchart-----Each Dose Group

| | Screening Stage | Cell Collection Stage | Baseline Stage [17] | Cell Transplantation Stage | Follow-up Stage | | | | | |
|---|---|---|---|---|---|---|---|---|---|---|
| **Content** | **14 days** | **1 day** | **0-5 days before transplantation** | **1 day** | **24 hours ± 4 hours after transplantation** | **1 week ± 2 days after transplantation** | **4 week ± 3 days after transplantation** | **12 week ± 5 days after transplantation** | **24 week ± 7 days after transplantation** | **Withdraw in advance [18]** |
| **Stage Code** | **V1** | **V2** | **V3** | **V4** | **V5** | **V6** | **V7** | **V8** | **V9** | |
| **Sign Informed Consent** | X | | | | | | | | | |
| **Check Inclusion/Exclusion Criteria** | X | | X | | | | | | | |
| **Demographic Characteristics [1]** | X | | | | | | | | | |
| **Medical History and Diagnosis [2]** | X | | | | | | | | | |
| **Symptoms and Physical Examination [3]** | X | X | X | X | X | X | X | X | X | X |
| **Vital Signs [4]** | X | X | X | X | X | X | X | X | X | X |
| **Blood Routine, Urine Routine [5]** | X | | X | | | X | X | X | X | X |
| **Blood Chorionic Gonadotropin Blood Test [6]** | X | | X | | | | | | X | X |
| **Blood Biochemistry Test [7]** | X | | X | | | X | X | X | X | X |
| **Arterial Blood Gas Test [8]** | | | X | | | | | X | X | X |

| | | | | | | | | | | |
|---|---|---|---|---|---|---|---|---|---|---|
| **Tetrachoric Assay of Blood Coagulation [9]** | X | | X | | | | | X | X | X |
| **Myocardial Enzyme Spectrum Assay [10]** | X | | | | | | | | X | X |
| **Autoantibody Test [11] (ANA, ENA, CCP, ANCA)** | X | | | | | | | | X | X |
| **Lung Cancer Markers [12]** | X | | | | | | | X | X | X |
| **Serological Examination for Syphilis, HIV, HBV, HCV [13]** | X | | | | | | | | X | X |
| **12-lead electrocardiogram [14]** | X | | X | | | X | X | | X | X |
| **Chest HRCT** | X | | | | | | | X | X | X |
| **Lung Function (Ventilation, Diffusion) [15]** | X | | X | | | | X | X | X | X |
| **Cell Collection through Bronchoscopy** | | X | | | | | | | | |
| **Cell Therapy through Bronchoscopy** | | | | X | | | | | | |
| **6 Minute Walking Distance** | | | X | | | | X | X | X | X |
| **SGRQ** | | | X | | | | X | X | X | X |
| **mMRC** | X | X | X | | | | X | X | X | X |
| **Acute IPF Exacerbation 16** | X | X | X | X | X | X | X | X | X | X |
| **Concomitant Medication** | X | X | X | X | X | X | X | X | X | X |

| | | | | | | | | | | |
|---|---|---|---|---|---|---|---|---|---|---|
| **Adverse Events** | | X | X | X | X | X | X | X | X | X |
| **Severe Adverse Events** | | X | X | X | X | X | X | X | X | X |

#### Note:

1. Including date of birth, gender, and ethnic.

2. Medical history: history of the present illness, past medical history, history of food and drug allergies, smoking history, drinking history, drug history, history of substance abuse.

3. Physical examination: skin mucous membrane, lymph nodes, head and neck, chest, abdomen, spine and limbs, musculoskeletal system, nervous system.

4. Vital signs: blood pressure, pulse, breathe, body temperature.

5. Blood routine: white blood cell count (WBC), neutrophil count (Neu), lymphocyte count (Lym), monocyte count (Mon), eosinophil count (Eos), basophil count (Bas), red blood cell count (RBC), hemoglobin (HGB), hematocrit (HCT), mean corpuscular volume (MCV), mean corpuscular hemoglobin concentration (MCHC), platelet (PLT). Urine routine: urine glucose (U-GLU), ketone bodies (KET), urine bilirubin (BIL), urobilinogen (UBG), urine protein (PRO), urine latent blood (BLD), urine pH, urine specific gravity (SG), urine red blood cells (URBC), urine white blood cells (UWBC).

6. Human chorionic gonadotropin (HCG): only for female patients (unapplicable for those who have been menopausal for more than a year).

7. Blood biochemistry: total protein (total protein), albumin (ALB), total bilirubin (TBIL), direct bilirubin (DBIL), alanine aminotransferase (ALT), aspartate aminotransferase (AST), alkaline phosphatase (ALP), γ-glutamyl transpeptidase (GGT), lactate dehydrogenase (LDH), urea (UREA), creatinine (Cr), uric acid (URIC), creatine kinase (CK), blood glucose (Glu), potassium ($K^+$), sodium ($Na^+$), chloride ($Cl^-$), calcium ($Ca^+$).

8. Arterial blood gas test: pH, $PaCO_2$, $PaO_2$, $HCO_3$.

9. Blood coagulation test: international normalized ratio (INR), activated partial thromboplastin time (APTT).

10. Myocardial enzyme spectrum: creatine kinase (CK), creatine kinase isoenzymes MB (CK-MB), lactate dehydrogenase (LDH).

11. Autoantibody test: antinuclear antibodies (ANA), extractable nuclear antigen (ENA) antibodies, cyclic citrullinated peptide (CCP) antibodies, antineutrophil cytoplasmic antibodies (ANCA).

12. Lung cancer markers: cytokeratin 19 fragment (CYFRA21-1), neuron-specific enolase (NSE), squamous cell carcinoma (SCC) antigen, carcinoembryonic antigen (CEA), carbohydrate antigen 125 (CA125).

13. Serological examination for syphilis, HIV, HBV, HCV: Qualitative test for HBV first. If its outcome is positive, do a quantitative measure for HBV next.

14. 12-lead electrocardiogram: heart rate, PR interval, QRS complex, QT interval (uncorrected), corrected QT interval QTc (QT/RR1/2), and an overall comment (normal, abnormality without clinical significance, abnormality with clinical significance, need more interpretation). The original electrocardiogram with signature will be stored in research center, the results of electrocardiogram examination are recorded on CRF or eCRF.

15. Lung function (ventilation, diffusion): height, weight, forced vital capacity (FVC), forced expiratory volume in 1 second (FEV1), ratio of FEV1 to FVC, diffusing capacity of the lungs for carbon monoxide (DLCO), alveolar ventilation (VA), the ratio of diffusing capacity of the lungs for carbon monoxide to alveolar ventilation (DLCO/VA). DLCO is measured with single-breath method. The examination results within 3 months before cell therapy can be accepted as the lung function test result at the baseline stage.

16. Idiopathic pulmonary fibrosis acute exacerbation: an acute, clinically significant respiratory deterioration without clear cause, acute worsening of dyspnea and decreased lung function, leading to respiratory failure or even death.

17. Patients should be checked again for inclusion at baseline stage. Results of myocardial enzyme spectrum, autoantibody test, lung cancer markers, serological examination for syphilis, HIV, HBV, HCV, and chest HRCT at the screening stage can be accepted.

18. If abnormality with clinical significance was reported when the patient withdraws in advance, the patient should be followed up until the recovery from the adverse events, reaching a stable status or other clear outcomes as required by the investigator

#### 1.5 FOLLOW-UP PLAN

All follow-up visits during the trial: V5 (24 hours ± 4 hours after transplantation), V6 (1 week ± 2 days after transplantation), V7 (4 weeks ± 3 days after transplantation), V8 (12 weeks ± 5 days after transplantation), V9 (24 weeks ± 7 days after transplantation). During the study, the vital signs, laboratory examinations, 12-lead electrocardiogram, chest HRCT, pulmonary functions, 6-minute walking distance, questionnaires, concomitant medications, adverse events, etc. will be checked and recorded.

(1) After collecting lung tissue specimens through bronchofiberscope, keep the patient under close observation for 2 hours. The investigator will determine whether the observation period should be extended according to the tolerance of the patient;

(2) After cell transplantation through bronchofiberscope, keep the patient under close observation in the outpatient or hospital for 2-4 hours after administration, and the investigator will determine when the patients can leave based on the tolerance of the patients.

Safety follow-up stage after research: After the end of the follow-up stage in this study, the patient will enter the post-study safety follow-up until the patient’s death or lost. The patient will be followed up once each at 1, 2, 3, 5, 8, and 10 years after treatment, and then once every five years. The follow-up measures include lung function, chest HRCT, and survival. This is not included in the 24-week follow-up, for the patient's knowledge only.

#### 1.6 CRITERIA FOR DOSE ESCALATION TERMINATION

(1) Adverse events that occurred throughout the study period were assessed and graded according to CTCAE V5.0. Dose escalation should be terminated with one or more adverse events that were the same or similar and believed to be related (possibly, probably or definitely related) to the REGEND001 autologous therapy product reported on two or more patients of the same dose group after autologous transplantation:

1) hematological toxicity related to cell therapy

a) grade 4 neutropenia (ANC) for 3 days or more; or grade 3 and higher neutropenia with fever (ANC<1000/$mm^3$ with a single measure of oral temperature >38.3℃ or ≥38.0℃ for one hour);

b) grade 3 thrombocytopenia ($25\times10^9$/L ≤ platelet counting ＜ $50\times10^9$/L) with overt clinical bleeding symptoms, or grade 4 thrombocytopenia (with or without overt clinical bleeding symptoms);

c) other grade 4 hematological toxicity.

2)non-hematological toxicity related to cell therapy

grade 3 and higher non-hematological toxicity, except:

a) under medical intervention, grade 3 non-hematological toxicity lasting ≤ 72 hours (non-laboratory examination, such as nausea, vomiting, or diarrhea, etc.);

b) under medical intervention, grade 3 or 4 non-hematological toxicity of abnormal in laboratory examination lasting ≤ 7 days (laboratory examination, such as arrhythmia, cardiac insufficiency, and nervous system abnormalities);

c) other toxicity that is higher than baseline level and related to cell therapy, classified as clinically meaningful severe and/or unacceptable toxicity by investigator and sponsor.

(2) Acute IPF exacerbation and severe adverse events that are related (possibly, probably or definitely related) to REGEND001 cell therapy;

(3) Adverse events that making investigator and sponsor believe the dose should not be escalated anymore, regardless of the event grade or reason;

(4) If none toxicity that would terminate dose escalation happened under the highest 3.3 times standard dose, investigator and sponsor will discuss together regarding

whether dose escalation should continue; if discontinued, the study will stop.

##### 1.7 SAMPLE SIZE

This clinical research intends to enroll 15-24 idiopathic pulmonary fibrosis patients.

*If the patient drops out within 4 weeks after cell administration, additional patients should be supplemented in the corresponding dose group; if the drop out happens after 4 weeks, supplementary is not needed.

#### 2 SUBJECT SELECTION

##### 2.1 DIAGNOSIS CRITERIA

According to the guidelines for the diagnosis of idiopathic pulmonary fibrosis 2018 edition, the diagnosis of IPF needs to meet the following criteria:

(1) Ruling out other ILD with known reasons (such as exposure in living and working environments, connective tissue diseases and drugs), with the following second or third performance

(2) HRCT images with UIP features

(3) Meeting specific combinations of HRCT features and pulmonary pathology features for patients with lung disease.

#### 2.2 INCLUSION CRITERIA

(1) Male or female, aged between 50 to 75;

(2) Subjects diagnosed with IPF according to guidelines for the diagnosis of idiopathic pulmonary fibrosis 2018 edition;

(3) Subjects with 30%~79% of the predicted value in diffusing capacity for carbon monoxide (DLCO) and more than 50% of the predicted value in forced vital capacity (FVC) in pulmonary function tests 3 months before screening;

(4) Subjects with typical High-resolution computed tomography (HR-CT) imaging findings of idiopathic pulmonary fibrosis in the past 12 months;

(5) Subjects tolerant to bronchofiberscope;

(6) Subjects fully informed of the purpose, method and possible discomfort of the trial, agreeing to participate in the test, and voluntarily signing the informed consent;

(7) Subjects with good adherence, willingness to take medication and regular follow-up examinations as required by the protocol；

(8) Subjects able to understand and cooperate with the completion of pulmonary function tests.

#### 2.3 EXCLUSION CRITERIA

(1) Subjects who cannot tolerate cell therapy

(2) Pregnant or lactating women;

(3) Subjects with syphilis or any of human immunodeficiency virus (HIV), hepatitis B virus (HBV), hepatitis C virus (HCV) positive antibody; Of which stable HBV carriers after drug treatment (DNA titer ≤500 IU/mL or copy number <1000 copies/mL) and cured hepatitis C patients (HCV RNA is negative) can be enrolled；

(4) Subjects with malignant tumors or a history of malignant tumors;

(5) Subjects with taking drugs which caused lung fibroblast such as amiodarone in a long term before screening;

(6) Subjects with infections in lung or other site, including bacterial and viral infections, requiring intravenous treatment before cell transplantation;

(7) Subjects with a history of invasive or noninvasive mechanical ventilation within 4 weeks;

(8) Subjects with any of the following lung diseases: asthma, active tuberculosis, pulmonary embolism, pneumothorax, pulmonary hypertension, pneumoconiosis, etc.; lung cancer, bronchiolitis obliterans or other active lung

disease; Pneumonia currently or within the last 4 weeks; Pneumonectomy Previously;

(9) Subjects needing oxygen therapy currently (oxygen therapy time> 15h/d);

(10) Subjects suffering from serious other systemic diseases, such as myocardial infarction, unstable angina, liver cirrhosis, acute glomerulonephritis, connective tissue disease, etc.;

(11) Subjects with following results: leukopenia (leukopenia < 4×10^9/L) or agranulocytosis (leukocyte < 1.5×10^9/L or neutrophils < 0.5×10^9/L) of any cause; Blood creatinine > 2.5 times the upper limit of normal; Alanine transaminase (ALT) and Aspartate transaminase (AST) > 2.5 times the upper limit of normal values in the laboratory tests.

(12) Subjects with a history of mental illness or suicide risk, epilepsy or other central nervous system disorders

(13) Subjects with severe arrhythmias (such as ventricular tachycardia, frequent supraventricular tachycardia, atrial fibrillation, atrial flutter, etc.) or atrioventricular block of degree II or above, shown by 12-lead Electrocardiogram (ECG);

(14) Subjects with a history of abusing alcohol and illicit drug;

(15) Subjects who are allergic to cattle products;

(16) Subjects who participated in other clinical trials in the past 3 months;

(17) Subjects with poor compliance and difficult to complete the investigation;

(18) Investigators, employees of research centers or family members of them (none of whom are suitable to participate in the trial to ensure the objectivity of the research);

(19) Subjects who had an acute IPF exacerbation or hospitalized for other respiratory diseases 3 or more times in the past 1 year;

(20) Subjects who take nintedanib for medication within 1 month, or plan to continue taking nintedanib for medication;

(21) Subjects with other acquired or congenital immunodeficiency disorders, or with a history of organ transplantation or cell transplant therapy;

(22) Subjects whose expected survival may be less than one year judged by the investigator;

(23) Male participants of childbearing potential and female participants within childbearing age were reluctant to use effective contraception from the time of signing the informed consent to 6 months after cell therapy;

(24) Subjects assessed as inappropriate to participate in this clinical trial by investigator.

#### 2.4 WITHDRAW CRITERIA

##### 2.4.1 WITHDRAW DECIDED BY INVESTIGATOR

(1) Patients with bad adherence, impacting the safety and tolerance evaluation;

(2) Patients undergo adverse events, making the investigator believe further study is not suitable;

(3) Other situations of the patient making the investigator believe further study is not suitable.

##### 2.4.2 WITHDRAW DECIDED BY PATIENT

(1) Patients have the right to withdraw for any reason;

(2) Even though the patient has not officially withdrawn, follow-up, cell administration, or examinations were not conducted successfully as required by the protocol.

If the patient withdraws before finishing the study, the withdraw reason should be recorded in CRF and original note.

### 3 THE TESTED DRUG IN EXPERIMENT GROUP

#### 3.1 NAME

Name: REGEND001 autologous therapy product (REGEND001 is a commercial cell produced by expanded human bronchial basal cells)

Dose: $0.6\times10^6$/Kg/person-$3.3\times10^6$/Kg/person

#### 3.2 DRUG APPLICATION METHOD

##### 3.2.1 THE COLLECTION, ISOLATION, AND EXPANSION OF REGEND001 CELLS BEFORE TREATMENT

Based on CT images, two healthy lung segments between 3-5$^{th}$ level bronchi were identified, from where tissue specimens were collected through bronchoscopy. The sample brush (with a trace amounts of bronchial tissues) was put in a sterile tube, and transported to Jiangxi Xlotus Medical Science and Technology Co., Ltd for isolation of bronchia basal cells. The tissue sample was collected by PBS washing and prepared single-cell suspension by enzyme digestion for cell culture. The basal cell culture system is a patented technology granted by Regend Therapeutics to Jiangxi Xlotus Medical Science and Technology Co., Ltd, including a combination of special growth factors and composite material for mimicking basal layer environment in vitro, so bronchial basal cells can be expanded selectively while other mature epithelial cells and fibroblasts apoptosis by nature because they can't grow. After the expansion of bronchial basal cells for a certain period, it can be stored in liquid nitrogen as cell bank for a long term. Before the application to the patients, REGEND001 cells need to undergo a series of strict tests.

##### 3.2.4 THE DETAILED PROCEDURE OF REGEND001 CELL THERAPY

The key steps for the transplantation of REGEND001 autologous therapy product:

(1) The site of infusion includes middle lobe of right lung, lingula of left lung, basal segments in the lower lobes of both lungs. The upper lobes of both lungs are not infused;

(2) Infuse REGEND001 cells to each lung segments through bronchofiberscope, with 3 ml at each segment;

(3) Taking 3 ml of infusion product with 10 ml syringe, with 5 ml air inside to push infusion product to the distal lung at the speed of 4-6 s/ml;

(4) The cells take around 1.5 hours to establish close adhesion with the inflammatory and injured lung area. After transplantation, patients were required to assume a supine position for 2 hours;

(5) The patients are prohibited from eating and drinking for 2 hours, minimizing coughing and oral codeine is given if necessary.

#### 3.3 TREATMENT COURSE

REGEND001 autologous therapy product is planned for one administration, and the treatment is divided into two stages. First, the collection, isolation and expansion of bronchial basal cells; second, the transplantation of REGEND001 autologous therapy. The whole procedure takes around 4 to 8 weeks. After that, the patients will be followed up for 24 weeks. The patients need to undergo corresponding examinations at hospital, including safety examination and efficacy examination.

#### 3.4 CONCOMITANT MEDICATION

IPF drugs used by the patients, except nintedanib, are compatible with this clinical trial. The details of all concomitant medications should be recorded on the Concomitant Drug Use page of the Case Report Form, clarifying the reason for medication/treatment, administration/treatment methods, and start and end dates.

List of common IPF drugs used in clinic:

| No. | Drug | Recommended dose | Main effect | Note |
|---|---|---|---|---|
| 1 | Pirfenidone | 600～1800mg/d | Antiinflammation, antifibrosis, antioxidant | |
| 2 | Omeprazole | 20～60mg/d | Treating gastroesophageal reflux disease associated with IPF | |
| 3 | N－Acetyl Cysteine | 2～12mL/d | Antioxidants, relieve coughing | |
| 4 | Methylprednisolon | 500～1000mg/d | Acute IPF exacerbation, hormone | |
| 5 | Prednisolone | ≥1mg/kg/d | Acute IPF exacerbation, hormone | |

List of other compatible concomitant drugs in this clinical trial:

| No | Drug | Recommended | Main effect | Note |
|---|---|---|---|---|
| 1 | Moxifloxacin | 400～800mg/d | Treating respiratory tract | |
| 2 | Eucalyptol, limonene and pinene enteric soft | 600～1200mg/d | An expectorant that can dissolve the mucus | |
| 3 | Compound methoxyphenaine capsule | 3～6 capsules/d | antispasmodic | |
| 4 | Prednisolone | 10～60mg/d | Anti-inflammation, anti-allergy | |
| 5 | Alprazolam | | Improve sleeping | |

#### 3.5 PACKAGING AND LABELING

##### 3.5.1 PACKAGING SPECIFICATIONS

14 mL per package (Prior to product release, the required dosage is prepared directly in the production area, for example, 3 bags totaling 42 ml).

##### 3.5.2 PACKAGING REQUIREMENTS

The product will be packaged in sealed sterile bags.

##### 3.5.3 LABELS

a. Inner packaging label

8cm×6cm

#### 3.6 CELL ALLOCATION

The cells will be strictly used for autologous treatment. Once the cells are prepared, they will be immediately transported to the clinical facility by a designated individual and received by responsible personnel. The cells will be used by the managing physician of the subject.

#### 3.7 PRODUCT STORAGE AND APPLICATION

Storage conditions: 2-8°C

Cell Administration: REGEND001 autologous cell suspension is delivered into the distal airways using a fiberoptic bronchoscope.

#### 3.8 DRUG MANAGEMENT

A designated individual will be responsible for managing the cells. The cells will be counted after each use, and any unused cells and packaging will be collected and returned.

#### 3.9 CODE ESTABLISHMENT

The patient identification number will be consistent with the cell preparation number. The numbering will start from the patient's admission to the hospital, and all related documents will use the same numbering system.

#### 3.10 QUALITY ASSURANCE MEASURES FOR REGEND001 CELL TRANSPORTATION

(1) First, ensure that the real-time temperature recorder is in good condition (with sufficient battery and calibrated within the past year). Set the temperature alarm upper limit to 8.1°C and the lower limit to 1.9°C.

(2) The packaged REGEND001 autologous therapy product, including both primary and secondary packaging, should be placed in a temperature-controlled box equilibrated at 2-8°C for transportation. Install an opened real-time temperature recorder inside the temperature-controlled box to monitor the temperature throughout the journey. Set the recording interval of the temperature recorder to 15 minutes, ensuring uninterrupted temperature monitoring, continuous recording, data storage, and alarms for temperature deviations. If air transportation restricts the use of temperature recorders, confirm the temperature during transportation using corresponding temperature indicator strips.

(3) Upon arrival of the REGEND001 autologous therapy at the clinical institution, personnel should inspect the

integrity of the packaging and confirm that the temperature displayed on the temperature recorder or indicated by the temperature indicator strip is within 2-8°C. Transport personnel should promptly remove the temperature recorder to verify the temperature inside the temperature-controlled box throughout the transportation process and complete the relevant documentation.

### 4 CONTROL GROUP

The control group is not involved in the early trial.

### 5 STUDY PROCEDURES

All subjects must sign an informed consent form prior to screening.

The study physician will provide the subjects with complete and truthful information about the background, objectives, trial design, direct and indirect benefits that the subjects can obtain, potential risks, and any other relevant information related to this clinical trial, in a quiet and private environment. The subjects will be allowed to think independently and discuss with the physician, ask any questions they have, and receive help to fully understand all the information. After the subjects make a voluntary decision to participate, they and the study physician will sign the informed consent form simultaneously.

Please refer to the clinical trial flowchart at the beginning of the protocol for details on the specific study procedures.

#### 5.1 FIBEROPTIC BRONCHOSCOPY EXAMINATION

(1) Preoperative preparation: The patient's medical history, physical examination, CT examination, and laboratory tests should be thoroughly evaluated. The purpose and significance of the examination should be explained to the patient to eliminate any concerns. The patient should fast for at least 4 hours before the bronchoscopic procedure. Sedatives can be administered to those who are more anxious.

(2) Local anesthesia: 2% lidocaine spray anesthesia is used in combination with inhalation anesthesia until there is a sense of obstruction in the throat.

(3) Procedure steps: The patient is typically placed in a supine position, and the bronchoscope is inserted through the nostril. During the examination, the tracheal mucosal folds and cartilage rings should be observed for clarity. The mucosa of the bronchial tube should be smooth and the color normal, with no new growths,

ulcers, or bleeding points. The nature of any secretions and the sharpness of the interstitium should also be noted, and detailed records should be kept.

(4) Specimen collection: If there are any new growths, bronchoscopic biopsies should be performed.

(5) Postoperative: The patient should fast for 3 hours, and food and water should only be consumed after the anesthesia has worn off to avoid aspiration. The patient should be informed that there may be blood in their sputum post-procedure. If the patient develops a fever, antibiotic treatment should be administered. Oxygen should be administered to patients who experience shortness of breath or hypoxemia.

#### 5.2 MANAGEMENT OF INELIGIBLE SUBJECTS DURING CELL CULTURING

In the time interval between subject enrollment and cell treatment (usually 4-8 weeks), if a subject who initially met the inclusion criteria becomes unsuitable for cell therapy (e.g., acute exacerbation of the condition, occurrence of severe complications), the subject's physical condition will be assessed by the researchers. If it is anticipated that the subject's condition can meet the standards for receiving cell transplantation after relevant interventions and observations, the cell therapy preparation work will be temporarily suspended. In such cases, there is no need to obtain new cells, and the cells that have already been cultured or partially cultured will be cryopreserved according to the relevant regulations of the production facility. Once the subject is medically cleared, they can directly proceed to the trial, reschedule the treatment time, and use the cells for treatment after cell recovery, culturing, and testing. If, based on the researcher's assessment, the subject is deemed unsuitable to continue participating in the study (such as an anticipated lack of short-term improvement in the physical condition), the subject will be considered a screening failure, and their cells will be destroyed. Before the completion of the entire study, a subject can undergo a maximum of two baseline assessments to verify the inclusion criteria. If the subject fails to meet the criteria for cell therapy during the second baseline assessment, the subject will be withdrawn from the trial, and the cells will be destroyed or cryopreserved for long-term storage if requested by the subject. If, until the end of the study, the subject is unable to meet the inclusion criteria, the subject will be withdrawn from the study, and the cells will be destroyed or cryopreserved for long-term storage if requested by the subject.

Considerations regarding the standards or scope of pre-treatment examinations for subjects undergoing cell therapy primarily involve the researcher's further verification of the inclusion criteria. If the subject still meets the selection criteria and does not meet any exclusion criteria, and there are no special circumstances, the researcher will assess and determine that the subject is still suitable for receiving cell therapy, and the treatment will proceed as planned. If the subject does not meet the inclusion criteria or meets any exclusion criteria, the treatment will be temporarily suspended (with the already cultured cells cryopreserved for later treatment), for example, the subject's condition has made significant progress: the increased risk of bronchoscopy due to worsening pulmonary inflammation, a follow-up CT scan (due to disease requirements or treatment interval exceeding 8 weeks) reveals concurrent lung cancer with a life expectancy of less than 1 year and posing a high risk of frequent acute exacerbations after expected cell therapy. In such complex situations, after comprehensive evaluation, the researcher deems the subject unsuitable for receiving cell therapy, and these circumstances can be considered as unacceptable treatment criteria or scope.

#### 5.3 MANAGEMENT OF FAILED CELL CULTURES

In the pre-treatment cell culture process, if it is observed that cells cannot be cultured, exhibit abnormal proliferation, or fail to meet quality requirements during quality inspection, it is necessary to investigate the reasons. In such cases, the technical department will design appropriate measures and arrange for the subjects to provide new samples for re-culturing of cells. Cultured cells that do not meet the required standards will be disposed of according to relevant regulations.

If the re-culturing process is successful, the subjects will be included in the clinical trial and proceed as planned. However, if re-culturing fails again, the subject will be excluded from the clinical trial.

### 6 SAFETY ASSESSMENTS

- Primary Endpoints:

Incidence and severity of cell therapy-related adverse events (AE) within 24 weeks post-treatment.

- Secondary Endpoints:

Incidence of complications during bronchoscopy within 24 weeks post-treatment;

Changes in pulmonary tumor markers compared to baseline at 12- and 24-weeks post-treatment;

Changes in abnormal values of routine safety checks compared to baseline within 24 weeks post-treatment (blood routine, urine routine, blood biochemistry and electrolytes, electrocardiogram).

Frequency and severity of acute IPF exacerbation events within 24 weeks post-treatment.

Corresponding indicators will be tested during follow-up observation, as follows.

#### 6.1 PHYSICAL AND VITAL SIGNS EXAMINATION

Physical and vital signs examination including height, weight, medical history, etc.

#### 6.2 LABORATORY TESTS

- Blood routine: White Blood Cell Count (WBC), Neutrophil Count (Neu), Lymphocyte Count (Lym), Monocyte Count (Mon), Eosinophil Count (Eos), Basophil Count (Bas), Red Blood Cell Count (RBC), Hemoglobin (HGB), Hematocrit (HCT), Mean Corpuscular Volume (MCV), Mean Corpuscular Hemoglobin Concentration (MCHC), Platelet Count (PLT).
- Urine routine: Urine Glucose (U-GLU), Ketones (KET), Urine Bilirubin (BIL), Urinary Urobilinogen (UBG), Urine

Protein (PRO), Urine Blood (BLD), Urine pH, Urine Specific Gravity (SG), Urine Red Blood Cells (URBC), Urine White Blood Cells (UWBC).

- Blood biochemistry: Total Protein (TP), Albumin (ALB), Total Bilirubin (TBIL), Direct Bilirubin (DBIL), Alanine Aminotransferase (ALT), Aspartate Aminotransferase (AST), Alkaline Phosphatase (ALP), Gamma-Glutamyl Transferase (GGT), Lactate Dehydrogenase (LDH), Blood Urea Nitrogen (BUN), Creatinine (Cr), Uric Acid (URIC), Creatine Kinase (CK), Blood Glucose (Glu), Potassium Ion (K+), Sodium Ion (Na+), Chloride Ion (Cl-), Calcium Ion (Ca+).

#### 6.3 12-LEAD ELECTROCARDIOGRAM (ECG) EXAMINATION

The 12-lead ECG examination should record heart rate, rhythm, PQ or PR interval, QRS interval, QT interval (uncorrected), and QTc (QT/RR1/2) and give an overall evaluation (normal, clinically insignificant abnormality, clinically significant abnormality, need further explanation). The signed original ECG will be archived at the study center, and the ECG examination result will be recorded on the CRF or eCRF.

#### 6.4 BLOOD GAS ANALYSIS

PH，$PaCO_2$，$PaO_2$，$HCO_3$

#### 6.5 PULMONARY TUMOR MARKERS

The pulmonary tumor markers include the following: Cytokeratin 19 fragment (CYFRA21-1), Neuron-specific enolase (NSE), Squamous cell carcinoma antigen (SCC), Carcinoembryonic antigen (CEA), Carbohydrate Antigen CA125.

#### 6.6 COMPLICATIONS OF BRONCHOSCOPY

Common complications of bronchoscopy include anesthesia accidents, bleeding, pneumothorax, laryngospasm, hypoxemia, infection, post-procedural fever, and other unforeseen events.

#### 6.7 ACUTE IPF EXACERBATION

Acute IPF exacerbation refers to a rapid deterioration of respiratory symptoms, worsening dyspnea, and decline in lung function without any identifiable cause, leading to respiratory failure or even death.

The diagnostic criteria for acute IPF exacerbation are as follows:

- History of IPF, or current clinical, imaging, and/or histological findings consistent with the diagnosis of IPF. If the previous diagnosis was not IPF based on the diagnostic criteria, current imaging and/or lung tissue pathology should indicate usual interstitial pneumonia pattern.
- Exacerbation of dyspnea or worsening of lung function within the past 30 days, not explainable by other causes.
- High-resolution computed tomography (HRCT) of the chest showing bilateral reticular or honeycomb patterns consistent with usual interstitial pneumonia, along with new ground-glass opacities and/or consolidations. If there is no prior contrast-enhanced HRCT, "new pulmonary imaging findings" can be disregarded.
- Absence of evidence for lung infection in tracheal secretions or bronchoalveolar lavage, including routine bacterial, opportunistic pathogen, and common viral examinations.
- Exclusion of other causes, including left heart failure, pulmonary embolism, and acute lung injury caused by other reasons. Causes of acute lung injury may include sepsis, aspiration, trauma, reperfusion pulmonary edema, pulmonary contusion, fat embolism, inhalation injury,

cardiac bypass surgery, drug toxicity, acute pancreatitis, transfusion of blood products, and cell transplantation.

When clinical data is incomplete or does not meet all five diagnostic criteria mentioned above, it is defined as suspected acute IPF exacerbation. Histopathologically, it is typically characterized by the coexistence of usual interstitial pneumonia (UIP) and diffuse alveolar damage (DAD), which may present with organizing pneumonia and significant fibroblast foci. Acute exacerbation leads to a worsening of lung function in IPF subjects, shortened survival time, poor treatment response, and high mortality rate.

### 7 EFFICACY ASSESSMENTS

- Changes in lung diffusing capacity for single-breath carbon monoxide (DLCO-sb) compared to baseline at 4-, 12-, and 24-weeks post-treatment.
- Changes in Forced Vital Capacity (FVC) compared to baseline at 4-, 12-, and 24-weeks post-treatment.
- Changes in the ratio of diffusing capacity for carbon monoxide/ the alveolar volume (DLCO/VA) compared to baseline at 4-, 12-, and 24-weeks post-treatment.
- Changes in 6-minute-walk test (6MWT) compared to baseline at 4-, 12-, and 24-weeks post-treatment.

Specific procedures are as follows: In the respiratory outpatient department, select a well-lit, well-ventilated, temperature-controlled corridor with a hard, straight, and flat surface, with a distance of 30 meters for the round trip, and place prominent markers at the starting and ending points. Before the test, explain the purpose, methods, and relevant precautions to the subjects. Participants should wear comfortable clothing and shoes during the test and may use walking aids such as canes. Subjects can have a light meal before the test, but should avoid excessive physical activity and warm-up exercises within 2 hours prior to the test. Subjects should sit quietly for at least 10 minutes at the starting point before the test. During the test, the timer should be started, and the subjects should walk at their own pace based on their usual daily activities from the starting point, turn around at the 30-meter endpoint, and the number of cycles completed in each round trip should be recorded. The testing personnel should not walk with the subjects or use overtly suggestive verbal or non-verbal encouragement. If subjects experience fatigue, dizziness, chest pain, intolerable dyspnea, lower limb spasms, cold sweats, or pallor during the test, the test should be immediately stopped. At the end of the test, shout "Stop" and ask the subjects to stop moving, marking their position. Blood pressure and heart rate should be measured before and after the test.

- Changes in the St. George's Respiratory Questionnaire (SGRQ) scale compared to baseline at 4-, 12-, and 24-weeks post-treatment.

- Changes in imaging of lung by high-resolution computed tomography (HR-CT) compared to baseline at 24 weeks post-treatment.

The HRCT scoring will be evaluated by the leading investigator of the coordinating center. The scoring will primarily rely on visual subjective assessment of the overall extent of ground-glass opacities, reticular abnormalities, honeycombing, and traction bronchiectasis, based on the imaging, without considering the subjects' clinical data and lung function.

### 8 OUTCOME MEASURES

#### 8.1 PRIMARY OUTCOME MEASURES

Incidence and severity of the cell therapy-related adverse events (AEs) within 24 weeks after treatment.

#### 8.2 SECONDARY OUTCOME MEASURES

- Incidence of complication related to bronchoscopy within 24 weeks after treatment.

- Changes in lung diffusing capacity for single-breath carbon monoxide (DLCO-sb) compared to baseline at 4-, 12-, and 24-weeks post-treatment.
- Changes in Forced Vital Capacity (FVC) compared to baseline at 4-, 12-, and 24-weeks post-treatment.
- Changes in the ratio of diffusing capacity for carbon monoxide/ the alveolar volume (DLCO/VA) compared to baseline at 4-, 12-, and 24-weeks post-treatment.
- Changes in 6-minute-walk test (6MWT) compared to baseline at 4-, 12-, and 24-weeks post-treatment.
- Changes in the St. George's Respiratory Questionnaire (SGRQ) scale compared to baseline at 4-, 12-, and 24-weeks post-treatment.
- Changes in imaging of lung by high-resolution computed tomography (HR-CT) compared to baseline at 24 weeks post-treatment.
- Frequency and severity of IPF exacerbation events within 24 weeks after treatment.
- Number of cases of participants with abnormal Blood routine results within 24 weeks post-treatment.
- Number of cases of participants with abnormal Urine routine results within 24 weeks post-treatment.

- Number of cases of participants with abnormal Blood biochemistry results within 24 weeks post-treatment.

- Number of cases of participants with abnormal 12-lead Electrocardiogram (ECG) results within 24 weeks post-treatment.

- Changes in tumor markers (Cytokeratin 19 fragment (CYFRA21-1), Neuron-specific enolase (NSE), Squamous cell carcinoma antigen (SCC), Carcinoembryonic antigen (CEA)) compared to baseline at 4-, 12-, and 24-weeks post-treatment.

### 9 CRITERIA FOR STUDY TERMINATION/DISCONTINUATION

The trial may be prematurely halted or terminated if there are sufficient reasons, including but not limited to the following:

(1) Occurrence of toxic reactions that meet the criteria for terminating dose escalation during the trial.

(2) Insufficient funding, patent disputes, or changes in national pharmaceutical development policies by the sponsor.

(3) Regulatory authorities or ethics committees request the suspension or termination of an approved trial.

In addition, the sponsor reserves the right to suspend or terminate the trial at any time.

Once the issues causing the trial suspension or termination, such as drug safety or protocol non-compliance, are resolved and approved by the sponsor, ethics committees, and regulatory authorities, the trial may resume.

After deciding to suspend or terminate the trial, any party involved (including but not limited to the sponsor, investigators, ethics committees, and regulatory authorities) should immediately issue a written notification to the other parties, providing the relevant reasons.

#### 10 FOLLOW-UP PLAN

During the trial, follow-up visits will be conducted as follows: V5 at 24 hours ± 4 hours after treatment, V6 at 1 week (±2 days) after treatment, V7 at 4 weeks (±3 days) after treatment, V8 at 12 weeks (±5 days) after treatment, and V9 at 24 weeks (±7 days) after treatment. Observations and recordings of vital signs, laboratory tests, electrocardiograms, chest high-resolution computed tomography (HRCT), pulmonary function, 6-minute walk distance, scale ratings, concomitant medications, adverse events, etc., will be conducted according to the study protocol.

After the completion of the follow-up period in this study, subjects will enter the post-study safety follow-up period until death or loss to follow-up. Follow-up visits will be conducted at

1, 2, 3, 5, 8, and 10 years after treatment completion, and subsequently, every 5 years.

### 11 ADVERSE EVENTS

#### 11.1 DEFINITION OF ADVERSE EVENTS

Adverse Events (AE): Refers to all adverse medical events that occur in subjects after receiving investigational drug. These events can manifest as symptoms, signs, diseases, or abnormal laboratory findings, but may not necessarily have a causal relationship with the investigational drug. They include the following situations:

(1) Exacerbation of pre-existing medical conditions/diseases (including worsening of symptoms, signs, and laboratory abnormalities) that were present before entering the clinical trial.

(2) Newly occurring AEs: Any newly occurring adverse medical condition (including symptoms, signs, and newly diagnosed diseases).

(3) Clinically significant abnormal laboratory values or results that are not caused by accompanying diseases.

Adverse Drug Reactions (ADR): Refers to any harmful or unexpected reactions related to the investigational drug that occur during the clinical trial. There must be at least a reasonable possibility of a causal relationship between the investigational drug and the adverse reactions, which means that the correlation cannot be ruled out.

Significant Adverse Events: Refers to any AE, other than Serious Adverse Events (SAE), that requires specific medical interventions (such as drug discontinuation, dose reduction, and symptomatic treatment), and results in significant hematological or other laboratory abnormalities.

#### 11.2 COLLECTION AND RECORDING OF ADVERSE EVENTS

Monitoring of Adverse Events (AE) will be conducted throughout the entire study, and it is the responsibility of the investigators to record all AEs observed during the study period. Any adverse medical events occurring from the initiation of autologous cell collection until the end of follow-up, regardless of severity or causal relationship with the investigational drug, must be documented in the medical records. Detailed documentation of the following information is required for all AEs:

- AE Description: Describe the AE using medical terminology rather than the language reported by the subjects. If applicable, include symptoms, signs, abnormal laboratory findings, and diagnosis. If the same AE occurs more than once in a subject and the subject has recovered between the two events, each occurrence should be recorded separately.

- AE Onset Date: The date when the subject first experienced the AE or when symptoms related to the AE appeared. If the AE is a clinically significant abnormal laboratory finding or test result, the date of sampling should be recorded.
- AE Outcome Date: The date of resolution of the AE or when symptoms related to the AE improved. If the AE is still ongoing, the outcome date does not need to be filled.
- Severity of AE.
- Causal Relationship between AE and the investigational drug.
- Actions Taken: (1) Actions taken with regard to the investigational drug: continued use, discontinued use, treatment completed, not applicable; (2) Actions taken regarding the AE: none, medication treatment, non-drug treatment.
- Outcome of AE: Disappeared, recovery with sequelae, improvement, stable, aggravated, death, subject refusal for follow-up or lost to follow-up, unknow.

#### 11.3 ADVERSE EVENT SEVERITY GRADING CRITERIA

The severity of Adverse Events (AE) in this study will be assessed according to the Common Terminology Criteria for Adverse Events (CTCAE) version 5.0. The grading of AE severity is as follows:

- Grade 1: Mild; asymptomatic or mild; clinical or diagnostic observations only; no treatment required.
- Grade 2: Moderate; requires minor, local, or non-invasive intervention; limitations in instrumental activities of daily living comparable to age-appropriate activities (instrumental activities of daily living refer to tasks such as cooking, shopping for clothes, using the telephone, managing finances, etc.).
- Grade 3: Severe or medically significant but not immediately life-threatening; results in hospitalization or prolonged hospital stay; causes disability; limitations in activities of daily living (activities of daily living include bathing, dressing, eating, personal hygiene, medication management, but not bedridden).
- Grade 4: Life-threatening; requires urgent intervention.
- Grade 5: AE-related death.

#### 11.4 ASSESSMENT OF ADVERSE EVENT (AE) CAUSALITY WITH INVESTIGATIONAL DRUG

The analysis of the association between adverse events and investigational drug should consider the following factors:

(1) The adverse event's temporal relationship to the administration of the investigational drug.

(2) Whether the adverse event disappears or diminishes after discontinuation or dose reduction of the investigational drug.

(3) Whether the adverse event reoccurs upon re-administration of the investigational drug.

(4) Whether the clinical or pathological manifestations of the adverse event are consistent with the known pharmacology and toxicology of the investigational drug or drug class.

(5) Whether the adverse event can be explained by the original disease or factors related to the subject or environment.

The association between adverse events and investigational drug can be categorized as follows:

(1) Definitely related: There is evidence of the use of the investigational drug. The occurrence of the adverse event has a reasonable temporal relationship to the administration of the investigational drug. It is supported by product labeling, similar drugs, or literature. The reaction

diminishes or disappears after discontinuation or dose reduction of the investigational drug, and reoccurs upon re-administration (if applicable). Other confounding factors, such as underlying diseases, have been excluded, and the explanation based on the investigational drug is more reasonable than other causes.

(2) Probably related: There is evidence of the use of the investigational drug. There is no history of re-administration, or although concomitant medications are present, the possibility of concomitant medication causing the adverse event is mostly ruled out.

(3) Possibly related: There is evidence of the use of the investigational drug. The occurrence of the adverse event has a reasonable temporal relationship to the administration of the investigational drug, supported by product labeling, similar drugs, or literature. It is unclear whether the adverse event reoccurs upon re-administration. Multiple drugs may have contributed to the adverse event, or disease progression factors cannot be excluded.

(4) Possibly unrelated: There is evidence of the use of the investigational drug. The occurrence of the adverse event has a reasonable temporal relationship to the administration of the investigational drug. It does not align with known adverse reactions in product labeling, similar drugs, or literature. It is unclear whether the adverse event

reoccurs upon re-administration. Other explanations may better account for the occurrence of the adverse event.

(5) Definitely unrelated: The investigational drug was not used, or there is evidence of its use but no temporal correlation with the occurrence of the adverse event, or there is a clear alternative cause for the adverse event.

### 12 SERIOUS ADVERSE EVENTS

#### 12.1 DEFINITION OF SERIOUS ADVERSE EVENTS

Serious Adverse Events (SAE): Refers to adverse medical events that occur in subjects following the administration of investigational drug, including death, life-threatening conditions, permanent or severe disabilities or functional impairments, subjects requiring hospitalization or prolonged hospital stays, as well as congenital abnormalities or birth defects.

The following circumstances are not considered SAE reports:

(1) Hospitalizations or prolongations of hospital stays resulting from adverse events in the clinical study are considered SAEs. However, the following hospitalizations are not included: rehabilitation facilities, nursing homes, routine emergency department admissions, and same-day surgeries (e.g., outpatient/day surgeries that do not involve

bed rest). Hospitalizations unrelated to the worsening of adverse events or prolongation of hospital stays, such as:

- Admission due to pre-existing conditions without the occurrence of new adverse events or worsening of pre-existing conditions (e.g., hospitalization for investigation of laboratory abnormalities present before the study).
- Hospitalization for administrative reasons (e.g., annual check-up).
- Hospitalizations specified by the study protocol during the clinical trial (e.g., operations performed according to protocol requirements).
- Elective hospitalizations unrelated to the worsening of adverse events (e.g., elective cosmetic surgery).
- Planned treatments or surgical procedures (to be documented in baseline data).
- Hospitalization solely for the use of blood products.

(2) Hospitalizations for administrative or social purposes, such as subjects admitted for recuperation or for insurance reimbursement purposes, are not considered AE reports.

(3) Diagnostic or therapeutic invasive procedures (e.g., surgery) and non-invasive operations should not be reported as AEs. However, if the condition leading to these

procedures meets the definition of an AE, it should be reported. For example, acute appendicitis occurring during the AE reporting period should be reported as an AE, while the appendectomy performed as a treatment for the AE should be documented as the therapeutic intervention for that AE.

(4) Hospitalization due to disease progression symptoms and signs during the trial should not be reported as SAEs. However, deaths resulting from disease progression, including its symptoms and signs, should be reported as SAEs.

#### 12.2 COLLECTION AND RECORDING OF SERIOUS ADVERSE EVENTS

This study should record serious adverse events (SAEs) from the start of autologous cell collection until the end of follow-up.

If a subject experiences a serious adverse event (SAE) during the trial, regardless of its relationship to the investigational drug, the investigator should immediately take appropriate treatment measures to ensure the subject's safety and promptly report to the clinical trial responsible person of the study sponsor. Upon learning of an SAE, the investigator must immediately inform the sponsor and/or Contract Research Organization (CRO), and the medical personnel of the sponsor and/or CRO should review the content of the SAE report form and provide feedback to the investigator. After multiple confirmations and reviews, the investigator signs and dates the report.

For SAEs with temporarily incomplete or uncertain information, it should still be reported to the sponsor and/or CRO personnel in a timely manner based on the principles of Good Clinical Practice (GCP), and supplementary reports should be provided in the form of follow-up reports once more information becomes available. The narrative section of the SAE report should provide detailed descriptions of the symptoms, severity, occurrence and management time, measures taken, follow-up time and method, as well as the outcome of the SAE. All SAEs should also be recorded in the Case Report Form (CRF), and the information provided in the SAE report form must be consistent with the data recorded in the CRF for the corresponding event.

If the investigator is unsure whether an adverse event (AE) qualifies as an SAE, it should be considered as an SAE until its nature can be determined. Such events should be promptly reported to the sponsor and/or CRO personnel in accordance with the principles of GCP.

Any deaths occurring during the trial, regardless of their relationship to the investigational drug, including deaths not related to the studied disease progression, require the investigator to provide timely rescue measures and submit necessary information, such as autopsy reports and final medical reports, to the sponsor and ethics committee.

For all SAEs, including those still in development at the end of the study until the last subject's follow-up, the investigator should submit follow-up reports to the sponsor. The reports should continue until the SAE is resolved, improved, stabilized, reasonably explained, or until the subject's death or loss to follow-up, ensuring that all issues are addressed. Detailed follow-up information should be provided, such as whether special treatment or hospitalization is required after the study. In the case of permanent damage, follow-up should continue until it is considered stable.

#### 12.3 ASSESSMENT OF CLINICAL LABORATORY TEST ABNORMALITIES AND OTHER ABNORMALITIES FOR ADVERSE EVENTS OR SERIOUS ADVERSE EVENT

Laboratory test abnormalities without clinical significance are not recorded as adverse events (AEs) or serious adverse events (SAEs). However, laboratory test abnormalities of clinical significance (referred to as "clinically significant abnormalities" or CS, such as clinical hematology and blood biochemistry) and evaluations of other abnormal situations (e.g., electrocardiograms, vital signs) should be assessed by the investigator. If they meet the definition of an AE or SAE, they must be recorded accordingly. Examples include:

- All laboratory test results that are clinically significant or meet the definition of an SAE.
- All laboratory test result abnormalities that require the subject to receive specialized routine treatment.

If the laboratory test abnormality is part of a syndrome, the syndrome or diagnostic result (e.g., anemia) should be recorded rather than the specific laboratory test result (e.g., decreased hemoglobin).

### 13 SUSPECTED UNEXPECTED SERIOUS ADVERSE REACTION (SUSAR)

#### 13.1 SUSAR DEFINITION

A SUSAR is defined as a serious adverse reaction with clinical manifestations that exceed the nature and severity described in the investigator's brochure, Summary of

Product Characteristics (SmPC), or other available information for investigational or marketed drugs.

The expectedness evaluation of a SUSAR report is based on the following four aspects:

(1) Whether the adverse event is explicitly stated in the investigator's brochure (IB).

(2) If the adverse event is listed in the IB, whether its severity is consistent with what is described (e.g., if the adverse event is described as mild in the IB, but the reported event results in hospitalization).

(3) If the adverse event is listed in the IB, whether its occurrence frequency is higher than what is stated.

(4) Other relevant factors that may contribute to the assessment.

#### 13.2 SUSAR REPORTING

All SUSARs must be promptly reported following the procedures outlined in the "Standards and Procedures for Rapid Reporting of Safety Data during Drug Clinical Trials" issued by the Center for Drug Evaluation (CDE) on April 27, 2018.

The applicant will medically evaluate the SAEs received, determine if they are SUSARs, and the SAE report forms identified as SUSARs will have their original data entered into the drug vigilance system by PV personnel. The data will undergo verification, questioning, MedDRA coding, and the generation of individual safety reports.

For SUSARs resulting in death or life-threatening situations, the sponsor is required to report to the CDE, the National Health Commission, all participating investigators and clinical trial institutions, and the ethics committee within 7 days of first notification. Follow-up information should be completed within the subsequent 8 days (counting the day of initial notification as day 0).

For non-fatal or non-life-threatening SUSARs, the sponsor should report to the CDE, the National Health Commission, all participating investigators and clinical trial institutions, and the ethics committee within 15 days of first notification. After the initial report, the sponsor should continue to monitor the SUSARs and provide timely follow-up reports on new information or changes to the previous report within 15 days of obtaining such information.

SAEs that occur after the completion of the clinical trial or the end of the follow-up period but before obtaining the evaluation and approval conclusion should be reported by the investigator to the sponsor. If they meet the criteria for SUSARs, they should also be promptly reported.

### 14 PREGNANCY

During the trial, if female subjects receiving investigational drugs or the partners of male subjects who receive investigational drugs become pregnant, the pregnancies will be recorded and reported to the ethics committee and the sponsor. Any female subject who becomes pregnant (intrauterine) during participation in this study must withdraw from the trial and immediately stop using the investigational drug. Any pregnancy events should be reported using the Clinical Trial Pregnancy Form.

For female subjects or the partners of male subjects who experience a pregnancy event, follow-up must be conducted until childbirth to determine pregnancy outcomes (including termination of pregnancy) and the maternal and infant conditions. Pregnancy complications and selective termination of pregnancy for medical reasons must be reported as AE or SAE, and spontaneous abortion must be reported as SAE.

Pregnancy events occurring in female subjects within 6 months after the end of the study treatment or events causing pregnancy in partners of male subjects within 6 months after the end of the study treatment should also be reported to the sponsor.

### 15 POTENTIAL RISK

#### 15.1 RISK OF BRONCHOSCOPY

Common complications associated with bronchoscopy include anesthesia accidents, bleeding, pneumothorax, laryngospasm, hypoxemia, infection, postoperative fever, and other unexpected events. Currently, clinical practice has matured and bronchoscopy is considered safe. This study only involves injection in the local pulmonary lobe, and the probability of complications is very low.

##### 15.1.1 MONITORING THE RISK OF ANESTHESIA-RELATED EVENTS

Monitoring the Risk of Anesthesia-related Events

(1) Anesthesiologists must enhance preoperative consultations, provide explanations to the subjects, develop appropriate anesthesia plans, and consult with senior physicians and department heads for difficult cases.

(2) Before performing anesthesia, check the functioning of anesthesia machines and monitoring devices. For any subject undergoing anesthesia, prepare for general anesthesia in advance, have endotracheal intubation equipment and corresponding rescue medications ready, and ensure that emergency supplies are readily available.

(3) Closely observe changes in the subject's vital signs and make accurate judgments and timely interventions. Adhere to a double-check system, properly label all medications used during anesthesia, and keep empty vials for verification until the subject leaves the operating room. Once anesthesia begins, the anesthesiologist should not leave the subject unattended, and severe penalties should be imposed for unauthorized departures. Select anesthesia medications based on specific circumstances, ensuring that the dosage remains within the prescribed range and adhering strictly to anesthesia operating procedures. Implement all preventive measures effectively. For resident physicians and trainee doctors, supervision should be rigorous, and difficult and critical cases should be handled by attending physicians.

(4) When performing combined anesthesia via intravenous and inhalation routes, prepare induction drugs in advance, secure any loose teeth of the subject appropriately, administer fluid replacement before induction, pay attention to the rate of medication administration and the subject's ventilation to avoid drastic fluctuations in blood pressure and inadequate ventilation. During endotracheal intubation, ensure adequate muscle relaxation and perform gentle maneuvers to avoid unnecessary injuries. Monitor closely the subject's vital signs during the procedure, and promptly communicate with the surgeon in case of any abnormalities,

ruling out possible interference from the surgical procedure and maintaining stable vital signs of the subject. When encountering challenging medical conditions, consultation with senior physicians is necessary, and the responsibility of attending physicians should be strictly enforced.

(5) Postoperative follow-up must be conducted within 48 hours to promptly report any issues to the supervising physician for timely intervention.

##### 15.1.2 MANAGEMENT OF ANESTHESIA-RELATED COMPLICATIONS

(1) Anesthesia Drug Allergy: Prior to anesthesia, administer a small amount of drug spray to the throat. If a significant allergic reaction occurs, the anesthesia drug should not be used. For severe allergic reactions or cases of toxic side effects, symptomatic treatment should be administered immediately, such as using vasopressors. Atropine can be used for bradycardia, and immediate tracheostomy should be performed for cases of laryngeal edema causing airway obstruction. In cases of cardiac arrest, cardiopulmonary resuscitation should be initiated.

(2) Insufficient or Excessive Anesthesia Medication: Insufficient local anesthesia can cause coughing, vomiting, and discomfort in the subject, and increase the likelihood of injury during bronchoscopy. Excessive local anesthesia can increase the risk of cardiovascular and central nervous

system toxicity. The total dose of lidocaine for adults should be limited to 8.2 mg/kg, calculated based on a weight of 60 kg, and the dose of 2% lidocaine should not exceed 25 mL. Extra caution should be exercised for elderly subjects or those with impaired liver or heart function. When achieving the desired anesthetic effect, the amount of lidocaine injected through the bronchoscope should be minimized.

(3) Anesthesia Accidents: Life-threatening complications during bronchoscopy procedures are often related to preoperative medication and local anesthesia. The risk is further increased in elderly subjects or those with severe comorbidities such as cardiovascular disease, chronic lung disease, liver or kidney dysfunction, epilepsy, or other mental disorders. Mild sedation and analgesia using drugs to alleviate anxiety and increase the pain threshold can improve subject cooperation, expedite the procedure, and reduce harm to the subject. Short-acting benzodiazepines (such as midazolam) can induce anterograde amnesia and control hypertension, with a lower incidence of respiratory depression compared to propofol. Opioids (such as fentanyl) can raise the threshold for stimulus response and reduce the occurrence of reflex reactions. In the presence of underlying organ dysfunction, adjustments should be made to the doses of benzodiazepines, opioids, anticholinergics, and local anesthetics.

#### 15.1.3 OTHER RISKS ASSOCIATED WITH BRONCHOSCOP

##### 15.1.3.1 Cardiac Arrhythmias

The most common is tachycardia accompanied by elevated blood pressure, often related to the stimulation and hypoxia caused by the bronchoscope. However, these are mostly self-limiting and quickly return to normal after the procedure is stopped. Severe and clinically significant arrhythmias and cardiac arrest are more common in individuals with pre-existing severe organic heart disease.

Insufficient anesthesia, passage of the bronchoscope beyond the vocal cords, intense stimulation of the tracheobronchial tree, and hypoxia are closely associated with cardiac arrhythmias. It is recommended to routinely monitor electrocardiography, blood pressure, and SaO2 during bronchoscopy, and the bronchoscopy room should be equipped with resuscitation equipment.

##### 15.1.3.2 Laryngospasm or Laryngeal Edema

These are commonly observed in subjects with difficult intubation or insufficient anesthesia, and in most cases, symptoms are relieved after removal of the bronchoscope. However, upper airway injuries during the procedure can lead to life-threatening laryngospasm. Proficient bronchoscopy skills can reduce the incidence of laryngospasm.

Special attention should be given to subjects with pre-existing bronchospastic diseases, superior vena cava syndrome, or a history of angioedema. Severe cases of laryngospasm or laryngeal edema should be treated promptly with oxygen supplementation, antihistamines, or intravenous administration of glucocorticoids.

15.1.3.3 Severe Bronchospasm

This is commonly observed in subjects undergoing bronchoscopy during an acute exacerbation of bronchial asthma. The bronchoscope should be immediately removed, and appropriate treatment should be provided for the severe asthma attack. In general, bronchoscopy should be relatively contraindicated during an acute asthma exacerbation. If bronchoscopy is necessary due to the patient's condition, it is recommended to perform the procedure under general anesthesia and mechanical ventilation.

For this reason, subjects with asthma should be rigorously excluded before enrollment.

15.1.3.4 Postoperative Fever

Transient fever after bronchoscopy is common and generally does not require special treatment. However, if the fever persists and the chest X-ray shows progressive infiltrates, the subject should be treated with antibiotics.

The possibility of fever increases in elderly subjects and those with airway obstruction after bronchoscopic interventions.

For subjects with underlying valvular heart disease and risk factors for endocarditis, the American Heart Association recommends prophylactic antibiotic use during rigid bronchoscopy but not during flexible bronchoscopy. However, for subjects with a history of prosthetic valve replacement, surgical vascular shunts, or endocarditis, prophylactic antibiotic use is necessary.

15.1.3.5 Hypoxemia

Decreased arterial oxygen tension (PaO2) during bronchoscopy is common. PaO2 generally decreases by approximately 20 mm Hg (1 mm Hg = 0.133 kPa) during bronchoscopy. Therefore, continuous monitoring of oxygen saturation (SaO2) should be performed for all subjects undergoing bronchoscopy. If necessary, end-tidal carbon dioxide concentration should also be monitored. Routine oxygen supplementation should be provided during the procedure to maintain SaO2 > 90%. Subjects with reduced lung function and sedation still require oxygen supplementation after the examination.

Bronchoscopy performed under local anesthesia with sedation can result in decreased oxygen saturation or inadequate ventilation, accompanied by a significant increase in PaCO2. Subjects with underlying chronic lung diseases may experience severe hypoxemia, leading to cardiac arrhythmias and life-threatening situations.

15.1.3.6 Hemorrhage

Bleeding during or after bronchoscopy is common, but significant bleeding is uncommon.

Preoperative assessment for subjects at potential risk of bleeding: For certain subjects, such as those with uremia, immunosuppression, pulmonary arterial hypertension, liver disease, coagulation disorders, or thrombocytopenia, the risk of bleeding is higher. Subjects without risk factors for bleeding before undergoing routine bronchoscopy do not require routine preoperative coagulation function screening. Conversely, for subjects at potential risk of bleeding, platelet count, prothrombin time, and activated partial thromboplastin time should be checked before bronchoscopy.

Currently, for subjects scheduled for bronchoscopic biopsy, it is recommended to perform platelet count, prothrombin time, and activated partial thromboplastin time tests prior to the examination. For subjects at risk of bleeding, even if biopsy is not performed and only routine bronchoscopy is conducted, platelet count and/or prothrombin time should be checked before the procedure. Long-term use of antiplatelet drugs such as clopidogrel should be discontinued for 1 week prior to biopsy. For subjects taking oral anticoagulants such as warfarin sodium, discontinuation of the medication for at least 3 days before the examination or administration of low-dose vitamin K antagonists should be considered. In rare cases where subjects must continue anticoagulant therapy, their international normalized ratio should be reduced to <2.5, and heparin should be used.

15.1.3.7 Pneumothorax

The incidence of pneumothorax is much lower than that of bleeding. The risk of pneumothorax is not related to the size of the biopsy forceps. Routine postoperative expiratory chest X-ray should be performed. In most subjects, even if pneumothorax occurs, it is usually mild and does not require chest tube insertion for closed thoracic drainage. However, for subjects with hypoxia and/or tension pneumothorax, chest tube placement is necessary for closed thoracic drainage.

#### 15.2 OTHER POTENTIAL RISKS DURING THE REGEND001 CELL THERAPY PROCESS

Initial treatments conducted in class A tertiary hospital in Shanghai and Chongqing demonstrated good safety. However, due to the relatively small number of overall cases, potential risks or unexpected situations are hereby disclosed.

##### 15.2.1 MULTIPLE ORGAN DYSFUNCTION SYNDROME (MODS)

Due to the rich capillary network in the lungs, there is a possibility that REGEND001 cells, if present in excessive amounts in the bloodstream, may cause multiple organ dysfunction. However, the REGEND001 cell therapy in this project is administered via the trachea, which closely resembles human physiology and clinical practice. By directly reaching the lung interstitium, it bypasses the multiple barriers involved in intravenous administration, such as the endothelium, interstitium, and basement membrane, significantly reducing the risk of multi-organ distribution to less than 0.01% and minimizing the possibility of organ dysfunction.

##### 15.2.2 LOCAL TUMOR FORMATION

Currently, there have been no reports of tumor formation from REGEND001 cells in animal experiments or in the clinical research conducted earlier. The term "local tumor formation" mainly refers to tumors appearing in the bronchial airway, most likely caused by cellular accumulation, particularly from natural apoptosis. The treatment methods for intrabronchial tumors include fiberoptic bronchoscopy for intraluminal removal or surgical intervention. If the tumor is malignant, further chemotherapy or radiotherapy may be required after surgery.

##### 15.2.3 OTHER POTENTIAL COMPLICATIONS

No clinical study-related complications were observed during the previous clinical research of REGEND001 cell therapy. Common complications during REGEND001 cell therapy mainly arise from fiberoptic bronchoscopy procedures, which were elaborated in the preceding text. However, it is challenging to estimate other potential complications at present. When unknown complications occur, corresponding treatment principles usually involve symptomatic support, surgical interventions, and consultation with relevant departments.

##### 15.2.4 ALLOGENEIC TRANSPLANTATION

Existing operating techniques ensure that cells from one subject will not be transplanted into another subject's body. Measures include the use of QR code scanning technology during cell harvesting, transportation, and processing for unique batch numbering and separate production for each subject.

If allogeneic transplantation of REGEND001 cells is discovered early, whole-lung lavage can be performed to remove the cells. If discovered later, follow-up observation is sufficient as allogeneic REGEND001 cells are difficult to engraft and survive.

##### 15.2.5 ACUTE EXACERBATION OF DISEASE

Acute IPF exacerbation refers to a sudden deterioration of the condition, worsened breathlessness, and decreased lung function without any clear cause, leading to respiratory failure or even death.

The diagnostic criteria for acute IPF exacerbation are as follows:

(1) History of IPF, or current clinical, imaging, and/or histological findings consistent with the diagnosis of IPF. If the previous diagnosis was not IPF based on the diagnostic criteria, current imaging and/or lung tissue pathology should indicate usual interstitial pneumonia pattern.

(2) Exacerbation of dyspnea or worsening of lung function within the past 30 days, not explainable by other causes.

(3) High-resolution computed tomography (HRCT) of the chest showing bilateral reticular or honeycomb patterns consistent with usual interstitial pneumonia, along with new ground-glass opacities and/or consolidations. If there is no prior contrast-enhanced HRCT, "new pulmonary imaging findings" can be disregarded.

(4) Absence of evidence for lung infection in tracheal secretions or bronchoalveolar lavage, including routine bacterial, opportunistic pathogen, and common viral examinations.

(5) Exclusion of other causes, including left heart failure, pulmonary embolism, and acute lung injury caused by other reasons. Causes of acute lung injury may include sepsis, aspiration, trauma, reperfusion pulmonary edema, pulmonary contusion, fat embolism, inhalation injury, cardiac bypass surgery, drug toxicity, acute

pancreatitis, transfusion of blood products, and cell transplantation.

When clinical data is incomplete or does not meet all five diagnostic criteria mentioned above, it is defined as suspected acute IPF exacerbation. Histopathologically, it is typically characterized by the coexistence of usual interstitial pneumonia (UIP) and diffuse alveolar damage (DAD), which may present with organizing pneumonia and significant fibroblast foci. Acute exacerbation leads to a worsening of lung function in IPF subjects, shortened survival time, poor treatment response, and high mortality rate.

Acute exacerbation of IPF is severe, with a high mortality rate. Clinically, pulse corticosteroid therapy (methylprednisolone 500-1000 mg/day) or high-dose corticosteroid treatment (prednisone ≥1 $mg \cdot kg^{-1} \cdot day^{-1}$) is still used. Although there is currently no consensus on the dosage, administration route, and duration of corticosteroid use, treatment can also include immunosuppressants, such as cyclophosphamide and cyclosporine A. Oxygen therapy, mechanical ventilation, and symptomatic treatment are the main therapeutic measures for patients with acute exacerbation of IPF.

### 16 DATA MANAGEMENT

Electronic Data Capture (EDC) technology is increasingly being utilized in clinical trials. It differs from traditional paper-based data collection methods and offers advantages such as real-time data entry, immediate identification of data errors, accelerated research progress, and improved data quality. As a result, regulatory authorities in various countries encourage the adoption of EDC technology in clinical trials to ensure data quality. The main functions of EDC are as follows.

#### 16.1 ECRF CONSTRUCTION

EDC systems are capable of generating electronic case report forms (eCRFs) that comply with the clinical trial protocol.

#### 16.2 DATA STORAGE AND AUDIT TRAIL

Once data is input into the EDC system, it retains an audit trail for all data modifications, preventing deletion or modification of audit trails.

#### 16.3 LOGICAL CHECKS

One of the key advantages of EDC is its ability to perform real-time automatic logical checks on data entry, such as data value ranges and logical relationships. The specific checks are outlined in the data verification plan tailored to each clinical trial. EDC systems are equipped with logic check modules.

#### 16.4 DATA QUERY MANAGEMENT

EDC systems have modules for generating, issuing, and closing data queries. Authorized data managers and/or clinical monitors can issue data queries to clinical research sites. The sites are responsible for confirming, explaining, or correcting the queried data. Authorized data managers can decide whether to close the query based on the response, or re-query data that does not meet requirements. All data query records should be preserved for reference.

#### 16.5 SOURCE DATA VERIFICATION CONFIRMATION

Source data verification confirmation is essential for ensuring the authenticity and integrity of clinical data. Clinical monitors are responsible for conducting source data verification on data stored in the EDC system. Source data confirmation can be accomplished using the data query function in the system.

#### 16.6 ELECTRONIC SIGNATURES

EDC systems have electronic signature functionality for all electronic records that require electronic signatures, including the generation, modification, maintenance, archiving, recovery, or transmission of any form of electronic forms. Electronic signatures can be implemented using login passwords and system-generated authorization codes. The association and legal equivalence between electronic signatures and handwritten signatures should be declared and confirmed by authorized users before implementing electronic signatures, and authorized electronic signatures have the same legal effect as their written handwritten signatures.

#### 16.7 DATABASE LOCKING

EDC systems have a locking function to prevent clean data that has been verified or confirmed from being altered. Once clinical data cleaning is completed, the EDC system can lock the database.

#### 16.8 DATA STORAGE AND EXPORT

EDC systems can store, export, or convert data into formats that meet clinical trial audit requirements and drug evaluation requirements.

For more information on Electronic Data Capture (EDC), refer to the "Guiding Principles for Electronic Data Capture Technology in Drug Clinical Trials."

### 17 STATISTICAL ANALYSES

#### 17.1 SAMPLE SIZE DETERMINATION

The sample size for this study is planned to be between 15 to 24 cases.

#### 17.2 DEFINITION AND SELECTION OF ANALYSIS SETS

Full Analysis Set (FAS): This set includes all enrolled cases that have received the study drug. FAS is used for reporting demographic data and baseline characteristics.

Per-Protocol Set (PPS): The PPS comprises cases that meet the inclusion criteria, do not meet the exclusion criteria, and have completed the treatment protocol. In other words, it includes cases that adhered to the trial protocol, completed Case Report Forms (CRFs) or electronic Case Report Forms (eCRFs), and are suitable for analysis (PP analysis). PP analysis is primarily used for efficacy assessment.

Safety Set (SS): The SS consists of actual data with safety indicators recorded from the initiation of fiberoptic bronchial cell collection. Safety-related missing values should not be carried forward. The SS may exclude certain cases for evaluation, such as cases exceeding the age inclusion criteria, but it does not include cases where safety assessments cannot be made due to the use of prohibited drugs. SS is used for safety evaluation analysis.

#### 17.3 STATISTICAL METHODS

##### 17.3.1 PROPOSED STATISTICAL METHODS

Outliers: Statistical and professional analysis is used to determine whether to include outliers.

Handling of Missing Values in Primary Efficacy Endpoint Data: If a primary efficacy endpoint data point is missing for an individual subject, the method of imputation will be determined based on statistical and professional considerations. For subjects who drop out, the last observation carried forward (LOCF) method will be used.

Analysis of Incomplete Cases: Analyze the reasons for each drop-out case individually.

Descriptive Statistics: Report the mean, standard deviation, maximum value, minimum value, median, quartiles, and percentages of occurrences.

##### 17.3.2 SAFETY EVALUATION

Adverse events (AEs) will be recorded after the first bronchoscopy collection and following the second bronchoscopy treatment. The description of AEs after the first bronchoscopy collection and the analysis of AEs after the second bronchoscopy treatment are as follows:

(1) Calculation of the incidence and severity of cell therapy-related adverse events (AEs) for each group.

(2) Calculation of the incidence of bronchoscopy-related complications for each group.

(3) Calculation of the mean, standard deviation, median, and interquartile range for the dynamic changes of each lung tumor biomarker in all groups.

(4) Presentation of the cross-tabulation of routine safety check abnormalities (complete blood count, urinalysis, blood biochemistry, electrolytes, electrocardiogram) between baseline and each visit.

(5) Recording the frequency and severity of acute IPF exacerbations

##### 17.3.3 EFFICACY ENDPOINT ANALYSIS

Quantitative variables will be described using measures of central tendency and dispersion, such as mean, standard deviation, maximum, minimum, median, and interquartile range. Qualitative variables will be presented as percentages for each group.

##### 17.3.4 STATISTICAL PRESENTATION

The primary method of reporting will be through self-explanatory tables, which include a title, captions, sample sizes, and footnotes.

For repetitive measurement data, results will be presented in tables along with statistical graphs to enhance readability.

##### 17.3.5 STATISTICAL SOFTWARE

Statistical analyses will be performed using SAS 9.4 (or higher version) software.

#### 18 TRIAL MANAGEMENT

Relevant laws, regulations, and technical guidelines that can be referenced for this clinical study include ICH-GCP, GCP, the "Drug Registration Administrative

Measures," the "Administrative Measures for Stem Cell Clinical Research (Trial)," the "Technical Guidelines for Research and Evaluation of Cell Therapy Products (Trial)," the "Interpretation of the Supreme People's Court and the Supreme People's Procuratorate on Several Issues Concerning the Application of Law in Handling Criminal Cases of Fabrication of Application Materials for Drug and Medical Device Registration," and other related interpretation documents.

Both the sponsor and the researchers should diligently fulfill their clinical research responsibilities, comply with relevant laws and regulations for clinical research, adhere to general and specific requirements of clinical research, and conscientiously complete all tasks involved in the clinical study.

#### 18.1 SPONSOR

##### 18.1.1 CLINICAL RESEARCH QUALITY MANAGEMENT

The sponsor should regard the protection of the rights and interests of the subjects, ensuring their safety, and the authenticity and reliability of the trial results as the fundamental starting points of the clinical trial.

The sponsor should establish a quality management system for drug (cell) clinical trials that covers the entire process of the clinical trial, including the design, implementation, documentation, evaluation, result reporting, and archiving of documents. Quality management includes effective trial protocol design, methods and processes for data collection, and information gathering to make decisions on important issues in the clinical trial.

The methods for quality assurance and quality control in drug (cell) clinical trials should align with the inherent risks of the clinical trial and the importance of information collection. The sponsor should ensure the operability of all aspects of the quality system, and the trial process and data collection should not be overly complicated. The trial protocol, CRF, and other relevant documents should be clear, concise, and consistent.

The sponsor should assume management responsibilities for all issues related to the clinical trial.

Depending on the needs of the trial, a research and management team for the clinical trial project may be established to guide and supervise the implementation of the clinical trial. Communication within the research and management team should be timely. All levels of personnel within the research and management team should participate when the drug regulatory authorities conduct inspections.

##### 18.1.2 CLINICAL RESEARCH RISK MANAGEMENT

(1) The risks of key trial processes and data should be identified. These risks should be considered on two levels: the system level (e.g., facilities and equipment, SOPs, computerized systems, personnel, suppliers) and the clinical trial level (e.g., investigational drug, trial design, data collection, and recording).

(2) Risk assessment should consider the likelihood of errors occurring under current risk controls, the impact of such errors on protecting the rights and ensuring the safety of subjects, as well as the reliability of the data, and the extent to which such errors can be detected.

(3) The sponsor should identify risks that can be reduced or accepted. Risk mitigation measures should be reflected in the design and implementation of the protocol, monitoring plans, clearly defined contracts for various responsibilities, SOP compliance, and various training programs.

(4) The sponsor should periodically evaluate risk control measures in light of new knowledge and experience gained during the trial process to ensure the effectiveness and applicability of current quality management activities.

##### 18.1.3 CLINICAL RESEARCH RISK MANAGEMENT

The sponsor is responsible for developing, implementing, and promptly updating the SOPs related to quality assurance and quality control of clinical trials, ensuring that the conduct of the clinical trial, the generation, recording, and reporting of data comply with the trial protocol, this specification, and relevant laws and regulations.

For critical aspects of clinical research, the sponsor is responsible for developing detailed and clear SOPs, including the "Standard Operating Procedures for Tracheal Basal Cell Tissue Sampling," "Standard Operating Procedures for REGEND001 Autologous Therapy Product and Transplantation," and "Standard Operating Procedures for Cold Chain Transportation," among others. Before the clinical research begins, researchers and related staff must undergo training and assessment. Only those who pass the assessment and receive an operational certificate issued by the sponsor can perform cell brushing and cell therapy operations to ensure the quality of clinical research implementation.

The entire process of clinical trials and laboratory testing must strictly follow quality management SOPs. Each stage of data processing must have quality control to ensure all data is reliable and the data processing is correct.

The sponsor must sign contracts with the researchers, their medical institutions, and all relevant units participating in the clinical trial, clearly defining the responsibilities of each party.

The sponsor should specify in the contracts with all relevant parties that inspections by domestic and foreign drug regulatory authorities, monitoring, and audits by the sponsor can directly access the trial site to review source data, source documents, and reports.

##### 18.1.4 CONTRACT RESEARCH ORGANIZATION

The sponsor may delegate part or all of the work and tasks of the clinical trial to a CRO, but the sponsor remains ultimately responsible for the quality and reliability of the clinical trial data and should supervise the work undertaken by the CRO. The CRO should establish a clinical trial quality assurance system and implement quality assurance and quality control.

##### 18.1.5 CLINICAL RESEARCH EXPERTS AND STAFFING

The sponsor may engage qualified medical experts to provide consultation on medical issues related to the clinical trial. When necessary, medical experts from external organizations may be hired to provide guidance.

The sponsor may select qualified biostatisticians, clinical pharmacologists, and clinical physicians to participate in the trial, including designing trial protocols and CRFs, developing statistical analysis plans, analyzing data, and writing interim and final trial summary reports.

The sponsor should employ qualified personnel to supervise the implementation of the trial, data processing, data verification, statistical analysis, and trial summary report writing. Qualified personnel may include experienced clinical research project managers, clinical medical professionals, preventive medicine professionals, clinical pharmacy professionals, clinical pharmacology professionals, nursing professionals, and others.

##### 18.1.6 CLINICAL RESEARCH DATA PROCESSING

The electronic data management system utilized by the sponsor must undergo compliant system validation to ensure the integrity, accuracy, and reliability of trial data. It should meet predefined technical performance criteria and maintain a validated status throughout the entire trial process.

The sponsor is required to retain all clinical trial data, including any additional data obtained from participants. These data should be specifically retained by the sponsor as essential documents within the clinical trial. The essential documents retained by the sponsor must comply with the regulatory requirements set forth by drug regulatory authorities for managing essential documents in drug clinical trials.

Any transfer of ownership of trial data must adhere to the requirements outlined in relevant regulations.

##### 18.1.7 CLINICAL RESEARCH RECORD PRESERVATION

If the sponsor terminates the clinical trial for any reason, they should retain relevant essential documents for at least 5 years from the formal cessation or suspension of the clinical trial, or in accordance with relevant regulations governing essential document management.

The sponsor's essential documents should be retained for at least 2 years after the drug is approved for marketing, or for at least 5 years from the formal cessation or suspension of the clinical trial. The retention period for documents may be extended as required by the drug regulatory authorities or the sponsor's internal regulations.

The sponsor should inform the researchers and the medical institutions employing them in writing about the requirements for preserving trial records. Additionally, when the trial-related records are no longer needed, the sponsor should inform the researchers and the medical institutions employing them in writing.

##### 18.1.8 ACCESS TO CLINICAL RESEARCH RECORDS

The sponsor should explicitly state in the clinical trial protocol or written agreements that researchers and the medical institutions employing them allow the sponsor's monitors, auditors, ethics committee reviewers, and drug regulatory authority inspectors to directly access the source data and source documents related to the clinical trial.

The sponsor should verify that each subject has signed a qualified and compliant informed consent form. During the clinical trial, monitors, auditors, ethics committee reviewers, and drug regulatory authority inspectors may directly access the original medical records related to the subjects' participation in the drug clinical trial.

##### 18.1.9 CLINICAL RESEARCH MONITORING AND AUDITING

The sponsor appoints monitors. Monitors should receive appropriate training and possess sufficient scientific and clinical knowledge necessary for clinical trial monitoring. Monitors must hold qualification certificates.

The purpose of monitoring is to ensure the rights of subjects, ensure the accuracy and completeness of trial records and reports, and ensure compliance with approved protocols and relevant regulations. Monitoring should comply with relevant policy and regulatory requirements.

The sponsor should ensure the implementation of clinical trial monitoring. The sponsor should establish a systematic, prioritized, risk-based approach to monitoring clinical trials.

The sponsor appoints auditors. Auditors should have extensive experience in clinical research and possess sufficient scientific and clinical knowledge necessary for clinical trial monitoring.

##### 18.1.10 COMPENSATION FOR SUBJECTS AND RESEARCHERS

The sponsor should provide legal and economic insurance or guarantees related to the clinical trial to researchers and the medical institutions employing them within the scope prescribed by relevant laws and regulations. However, this does not include damages caused by the fault of the researchers or their employing medical institutions.

The sponsor must bear the expenses for diagnosis, treatment, and appropriate economic compensation for subjects who suffer harm or death related to the trial. The sponsor and the researchers should promptly pay compensation to the subjects.

The methods and procedures for compensation provided to subjects by the sponsor must comply with relevant laws and regulations.

##### 18.1.11 CLINICAL TRIAL CONTRACTS

The contract signed between the sponsor and the researchers and their employing medical institutions should clearly define the responsibilities, rights, and interests of all parties involved in the trial while avoiding potential conflicts of interest. The trial funding in the contract should be reasonable and in line with market principles, while also addressing any potential conflicts of interest.

The contract should include provisions for: compliance with this specification and other relevant laws and regulations governing drug clinical trials during the implementation of the trial; adherence to the protocol approved by the ethics committee through negotiation between the sponsor and the researchers; compliance with data recording and reporting procedures; agreement to monitoring, auditing, and inspections; preservation of essential trial-related documents and their retention period; and arrangements for publication of articles, among others. The sponsor and the researchers and their employing medical institutions should sign and confirm the contract.

##### 18.1.12 INFORMATION ON INVESTIGATIONAL DRUGS

When formulating the clinical trial protocol, the sponsor should have sufficient non-clinical safety and efficacy trial data supporting the investigational drug. Alternatively, safety and efficacy data on dosage route, dosage, and duration of administration from populations previously treated with the drug should support the trial.

When significant new information is obtained, the sponsor should promptly update the investigator's brochure.

##### 18.1.13 SAFETY INFORMATION FOR CLINICAL TRIALS

The sponsor is responsible for assessing the safety of trial drugs during the trial period. The sponsor should promptly notify the researchers and their employing medical institutions and the drug regulatory authorities of any issues discovered during the clinical trial that may affect subject safety, trial implementation, or alter the approved opinion of the ethics committee.

##### 18.1.14 REVISION OF THE INVESTIGATOR'S BROCHURE

The sponsor must establish a written procedure for revising the investigator's brochure, requiring the brochure to be reviewed and revised at least once a year during the trial period. Depending on the developmental stages of new drug clinical trials and the acquisition of relevant safety and efficacy data during the clinical trial process, the investigator's brochure may require multiple revisions. The sponsor should inform the researchers of any significant new information obtained before updating the investigator's brochure and communicate with the ethics committee and/or drug regulatory authorities as necessary.

The sponsor is responsible for updating the investigator's brochure and delivering it to the researchers promptly. The researchers are responsible for submitting the updated brochure to the ethics committee.

##### 18.1.15 SPONSOR CONTACT INFORMATION

Sponsor Name: Regend Therapeutics XLotus (Jiangxi) Co, Ltd.

Address: Building 8, Nanchang National Pharmaceutical International Innovation Park Joint Research Institute, 269 Aixi Lake North Road, High-tech Development Zone, Nanchang, Jiangxi Province

##### 18.1.16 CELL QUALITY CONTROL

The production and preparation of cells must be conducted in GMP workshops. The collection, separation, cultivation, and transportation of cells must strictly adhere to SOPs. For cell transportation, corresponding quality testing, outbound testing, and release reports must be provided. Particularly, temperature control reports during transportation must be provided to ensure the quality of returned cells.

#### 18.2 INVESTIGATORS

##### 18.2.1 QUALIFICATIONS OF INVESTIGATORS

(1) Investigators must hold a valid practicing qualification in their affiliated medical institution.They should possess the professional knowledge, training experience, and clinical trial experience required for conducting clinical trials.The latest curriculum vitae and relevant qualification documents should be provided to the applicant, ethics committee, and drug regulatory authority.

(2) Familiarity with the clinical trial protocol provided by the applicant, investigator's manual, and information related to trial drugs (cells).

(3) Familiarity with and adherence to this regulation and relevant laws and regulations regarding clinical trials.

(4) Investigators and their affiliated medical institutions should be open to inspections and audits organized by the applicant and inspections by drug regulatory authorities.

(5) A researcher's work division authorization form signed by the principal investigator must be retained.

##### 18.2.2 MEDICAL AND CLINICAL TRIAL RESOURCES

(1) Investigators should be able to enroll a sufficient number of eligible subjects within the time frame specified in the clinical trial contract.

(2) Investigators should ensure the completion of the clinical trial within the specified time frame in the trial protocol.

(3) During the conduct of the clinical trial, investigators have the authority to manage personnel involved in the trial, have access to the required medical facilities for the trial, and ensure their correct and safe use.

(4) Investigators and their affiliated medical institutions should ensure that all personnel participating

in the clinical trial fully understand the clinical trial protocol and trial drugs, clarify their roles and responsibilities in the trial, and ensure the authenticity, integrity, and accuracy of the clinical trial data.

(5) The principal investigator supervises all investigators in implementing the trial protocol and takes measures to implement quality management of the clinical trial.

##### 18.2.3 COMPLIANCE WITH TRIAL PROTOCOL

(1) Investigators and their affiliated medical institutions should conduct trials in accordance with the clinical trial protocol approved by the ethics committee.

(2) Except for urgent changes to reduce immediate harm to subjects or administrative changes related to clinical trial management, such as replacing monitors or phone numbers, investigators may not modify or deviate from the clinical trial protocol without the consent of the applicant and approval of the ethics committee.

(3) Any deviations from the approved trial protocol should be recorded and explained by the investigator or designated personnel.

(4) If investigators intend to modify the clinical trial protocol, they must obtain the consent of the applicant and submit it for review by the ethics committee, and report to the drug regulatory authority if necessary. In order to eliminate immediate harm to subjects, if the investigator deviates from or changes the trial protocol without approval from the ethics committee, they should promptly report to the ethics committee and the applicant, providing reasons and, if necessary, report to the drug regulatory authority.

(5) After subjects enter the clinical trial, investigators should control concomitant medications, especially the use of similar drugs. Medical institutions and principal investigators should take measures, in addition to enhancing investigator training, to document relevant clinical trial information in the subjects' paper and electronic medical records; they should also use information systems to restrict the use of prohibited drugs as specified in the protocol and provide reminders for drugs requiring caution. Subjects should be informed that if they seek treatment at another hospital during the trial, they should inform the attending physician of their

participation in the clinical trial, especially regarding medication.

##### 18.2.4 TRIAL RECORDS AND REPORTING

(1) The principal investigator should oversee data collection at the clinical trial site and the performance of all research personnel.

(2) Investigators should ensure that all clinical trial data are obtained from source documents and trial records, are accurate, complete, legible, and timely. Source data should be traceable, clear, contemporaneously recorded, original, accurate, and complete. Corrections to source data must be documented, not obscure the initial data, reasons for the corrected data should be explained when necessary.

(3) Investigators should complete and modify case report forms (CRF or eCRF) according to the guidelines provided by the applicant, ensuring that data in various CRFs and other reports are accurate, complete, clear, and timely. Data reported in CRFs should match the source documents, and any inconsistencies should be reasonably explained. Any modifications to data in CRFs

should be clearly identifiable, with modification trails retained, reasons explained when necessary, and signed and dated by the modifier.

Applicants should have written procedures to ensure that modifications to CRFs are necessary, documented, and approved by the investigator. Investigators should retain relevant records of modifications and corrections.

(4) Investigators and their affiliated medical institutions should properly maintain trial documents according to the requirements of "Essential Documents for Clinical Trials" and the drug regulatory authority.

(5) Essential trial documents should be retained for 2 years after the approval of trial drugs for marketing or 5 years after the termination of the clinical trial. Applicants should specify in the contract with investigators and their affiliated medical institutions the duration, cost, and handling after expiration of essential documents.

(6) Applicants and investigators and their affiliated medical institutions should clearly specify trial-related financial matters in the contract.

(7) Investigators and their affiliated medical institutions should cooperate with and provide necessary trial-related records as requested by inspectors, auditors, ethics committees, or drug regulatory authorities.

##### 18.2.5 TRIAL PROGRESS REPORTS AND FINAL REPORTS

(1) Investigators should submit annual reports of clinical trials to the ethics committee or provide progress reports as requested by the ethics committee.

(2) If there are changes in the medical institutions or an increase in risks to participating subjects during the trial, investigators should promptly report to the applicant and ethics committee in writing.

Upon completion of the trial, investigators should report to their affiliated medical institutions; investigators and their affiliated medical institutions should provide a summary of trial results to the ethics committee and relevant clinical trial reports to the applicant as required by the drug regulatory authority.

##### 18.2.6 INFORMED CONSENT OF SUBJECTS

During the informed consent process, researchers must comply with the regulatory requirements of the drug regulatory authority, adhere to the provisions of this guideline, and adhere to the ethical principles of the Helsinki Declaration.

During the clinical trial process, when researchers obtain new information that may affect the continued participation of subjects in the trial, they should promptly inform the subjects or their legally authorized representatives in writing, such as by providing an informed consent form.

All relevant new information that needs to be communicated to the subjects, after approval by the ethics committee, should require subjects to sign a new informed consent form. Newly enrolled subjects should sign an updated informed consent form and other written materials.

Researchers or any other research personnel are prohibited from using coercion, inducement, or any other improper means to influence subjects to participate or continue participating in the clinical trial.

Any written or verbal information related to the trial must not contain language that would cause subjects and their legally authorized representatives to waive their legal rights, nor contain language that would absolve the researchers or their affiliated medical institutions, sponsors, or their agents from their responsibilities.

Researchers or designated research personnel should fully inform subjects about all relevant aspects of the clinical trial, including written information and the approval opinions of the ethics committee. Subjects lacking the capacity to provide informed consent should have their legally authorized representatives act on their behalf.

Oral and written materials provided to subjects, such as informed consent forms, should be in plain and understandable language and expression, making it easy for subjects, their legally authorized representatives, and witnesses to understand.

Before signing the informed consent form, researchers or designated research personnel should give subjects or their legally authorized representatives sufficient time and opportunity to understand the details of the trial and to thoroughly answer any questions related to the trial that subjects or their legally authorized representatives may have.

Subjects or their legally authorized representatives, as well as the researchers executing the informed consent, should sign and date the informed consent form separately. Researchers should ensure that the signed informed consent form is compliant.

In cases where subjects or their legally authorized representatives lack reading ability during the informed consent process, a neutral witness should assist and witness the informed consent.

Researchers should provide detailed explanations of the content of the informed consent form and other written materials to subjects or their legally authorized representatives and witnesses. If subjects or their legally authorized representatives verbally agree to participate in the trial and sign the informed consent form, witnesses should also sign and date the informed consent form to certify that subjects or their legally authorized representatives have been accurately explained the informed consent form and other written materials by the researchers, have understood the relevant content, and have agreed to participate in the clinical trial.

When the legally authorized representative consents to informed consent on behalf of the subject, they should inform and assist the subject in understanding the relevant information about the clinical trial as much as possible and try to have the subject personally sign the informed consent form and date it.

When a subject participates in a non-therapeutic trial and there are no anticipated clinical benefits, the subject must sign and date the informed consent form.

In cases where informed consent cannot be obtained from the subject before participating in a clinical trial in emergency situations, consent must be obtained from the subject's legally authorized representative. If the subject is unable to be informed in advance, and their legally authorized representative is also absent, the method of selection of the subject should be clearly stated in the clinical trial protocol and/or other documents, and written approval from the ethics committee should be obtained. Efforts should be made to obtain informed consent from the subject or their legally authorized representative to continue participating in the clinical trial as soon as possible.

When the legally authorized representative consents to the participation of the subject in a non-therapeutic trial, the following conditions must be met: the clinical trial can only be conducted in subjects lacking the capacity to provide informed consent; the subject lacks the capacity to provide informed consent; the anticipated risks to the subject are low; the negative impact on the subject's health has been minimized, and the law does not prohibit the conduct of such trials; the inclusion of such subjects has been approved by the ethics committee.

Unless in exceptional circumstances, non-therapeutic trials can only be conducted in subjects with diseases or conditions for which the investigational drug is applicable. Subjects should be closely monitored during the trial, and if subjects exhibit excessive pain or discomfort, they should be withdrawn from the trial.

##### 18.2.7 CONFIDENTIALITY OF PARTICIPANT DATA

This study only collects and processes data from participants that are essential for studying the effectiveness, safety, quality, and application of cell drugs.

When collecting and using this data, confidentiality will be fully ensured and relevant laws and regulations protecting participant privacy will be complied with.

The primary investigator will ensure that:

(1) The process of collecting and processing data is fair and legal.

(2) The purpose of collecting data is specific, clear, and lawful; and the data will not be further processed in a manner inconsistent with these purposes.

(3) The data collected is sufficient and relevant to the research purposes and does not collect data unrelated to the research purposes.

(4) The collected data is accurate and, when necessary, updated to be the most current.

Before collecting personal data, consent will be obtained from participants.

Participants have the right to access personal data through researchers and may request correction of any errors or incomplete data. Requests of this nature will be appropriately responded to considering their content and purpose, the status of the trial, and relevant laws.

Appropriate technical and management measures must be taken to protect participant personal information from unauthorized access and disclosure, accidental or unlawful destruction, accidental loss, and alteration. Throughout the study, personnel from the sponsor who are authorized to view participant data must maintain confidentiality of the information viewed.

#### 18.3 USE AND PUBLICATION OF INFORMATION

All unpublished information provided by the sponsor to the researcher, including but not limited to information regarding drugs (cells), commercial information of the sponsor (such as patent status, formulation, production process, basic research data, previous clinical data, and prescription data), and any data generated from this study, are confidential and proprietary to the sponsor.

Researchers agree to maintain confidentiality of this information and only use it for the purpose of completing this study, and shall not use it for any other purpose without the written consent of the sponsor.

Researchers should understand that the information obtained in this study will be used by the sponsor for further development of drugs (cells), and therefore this information may be provided to other clinical researchers or regulatory authorities. Researchers are obligated to provide all data derived from this trial to the sponsor for use.

The results of the study will be presented in the form of a clinical research report, which includes data from all participating units in the trial. Any materials derived from the trial and containing data beneficial to copyright protection belong to the sponsor and should be identified as authored by the sponsor or the owner of the copyright. The sponsor has the right to publish these materials and information without the consent of the researchers. If researchers wish to publish information from the trial, the original manuscript must be provided to the sponsor for review 60 days prior to submission or presentation.

Abstracts, posters, or other promotional materials will be reviewed more expeditiously. If the sponsor has a written request, researchers should further delay the publication of such material for 60 days to allow the sponsor to prepare patent applications. In cases of scientific rigor or regulatory compliance issues, the sponsor will discuss these matters with the researchers. The sponsor will not mandate modifications to the scientific content and does not have the authority to withhold data. Researchers should consider the integrity of multicenter trials, and central data can only be published under the following circumstances: when an article integrating all center results has been published; 12 months after all trial centers have concluded, abandoned, or terminated; the sponsor has confirmed the non-publication of multicenter study results. The attribution of this research publication will be determined according to widely accepted standards in authoritative medical journals.

#### 18.4 PARTICIPANT INSURANCE

If participants suffer injuries or damages related to the study during the treatment period, compensation shall be made according to the relevant laws and regulations of the country. Detailed insurance provisions can be found in the informed consent form.

#### 19 EXPECTED PROGRESS AND COMPLETION TIME OF THE CLINICAL TRIAL

(1) Study Commencement: The implementation of this protocol will commence upon approval by the Ethics Committee.

(2) Mid-Clinical Coordination Meeting: The timing of this meeting will be determined based on the progress and completion status of the clinical study.

(3) Clinical Study Completion Time: The study is planned to be completed within 18 months after the intended start date.

(4) Data Collection, Statistical Analysis, and Summary Time: The clinical study summary will be completed within 6 months after the study is finished and the statistical analysis report is received.

#### 20 ETHICAL CONSIDERATIONS

This clinical study will adhere to the Helsinki Declaration (2008 version) and relevant regulations and guidelines for the management of cellular clinical research in China. Prior to the commencement of the clinical trial, the research center ethics committee must approve the study protocol.

Before each participant is enrolled in the study, the investigating physician is responsible for providing complete and comprehensive written information about the purpose, procedures, and potential risks of the study to the participant or their designated representative. Participants should be informed that they have the right to withdraw from the study at any time. Each participant must be provided with an informed consent form before being enrolled in the study. The investigating physician is responsible for ensuring that each participant signs the informed consent form before entering the clinical trial and keeping it in the study records.

### 21 MAIN REFERENCES

Hong KU, Reynolds SD, et al. Basal cells are multipotent progenitor capable of renewing the bronchial epithelium. American Journal of Pathology 2004. 164:577-588.

Zuo W, Zhang T, et al. p63+Krt5+ distal airway stem cells are essential for lung regeneration. Nature 2015 517,616-620.

Vaughan A, Brumwell, et al. Lineage-negative progenitor mobilize to regenerate lungepithelium after major injury. Nature 2015 517, 621-625.
